# An atlas of causal and mechanistic drivers of interpatient heterogeneity in glioma

**DOI:** 10.1101/2024.04.05.24305380

**Authors:** Serdar Turkarslan, Yu He, Parvinder Hothi, Carl Murie, Abran Nicolas, Kavya Kannan, James H Park, Min Pan, Alaa Awawda, Zachariah D Cole, Mark A Shapiro, Timothy J Stuhlmiller, Hwahyung Lee, Anoop P Patel, Charles Cobbs, Nitin S Baliga

**Author notes:** Corresponding author: Nitin S Baliga.

## Abstract

Grade IV glioma, formerly known as glioblastoma multiforme (GBM) is the most aggressive and lethal type of brain tumor, and its treatment remains challenging in part due to extensive interpatient heterogeneity in disease driving mechanisms and lack of prognostic and predictive biomarkers. Using mechanistic inference of node-edge relationship (MINER), we have analyzed multiomics profiles from 516 patients and constructed an atlas of causal and mechanistic drivers of interpatient heterogeneity in GBM (gbmMINER). The atlas has delineated how 30 driver mutations act in a combinatorial scheme to causally influence a network of regulators (306 transcription factors and 73 miRNAs) of 179 transcriptional “programs”, influencing disease progression in patients across 23 disease states. Through extensive testing on independent patient cohorts, we share evidence that a machine learning model trained on activity profiles of programs within gbmMINER significantly augments risk stratification, identifying patients who are super-responders to standard of care and those that would benefit from 2^nd^ line treatments. In addition to providing mechanistic hypotheses regarding disease prognosis, the activity of programs containing targets of 2^nd^ line treatments accurately predicted efficacy of 28 drugs in killing glioma stem-like cells from 43 patients. Our findings demonstrate that interpatient heterogeneity manifests from differential activities of transcriptional programs, providing actionable strategies for mechanistically characterizing GBM from a systems perspective and developing better prognostic and predictive biomarkers for personalized medicine.

## INTRODUCTION

Glioblastoma multiforme (GBM) is the most commonly diagnosed malignant brain tumor, with an incidence of 5 per 100,000 people in North America^1^. GBM is among the most difficult tumor types to treat^2^. Complete tumor resection is only rarely possible due to the highly invasive nature of GBM cells, which spread to the surrounding brain tissue. The current standard of care for GBM includes surgical resection, radiotherapy, and concomitant and maintenance temozolomide (TMZ) chemotherapy^3,4^. The prognosis of GBM patients who receive standard of care remains dismal, with a median survival time of approximately 15 months, a five-year survival rate of less than 10%, and a recurrence rate of ∼90%^5^. In a randomized Phase III clinical trial, GBM patients treated with radiation, TMZ, and tumor treating fields (TTF) had a median overall survival (mOS) of 20.5 months, compared to 15.6 months for patients receiving standard treatment without TTF^6^. Despite the marginal improvements, GBM tumors remain difficult to treat with a poor prognosis^2^.

One major challenge for the lack of clinical improvement of GBM tumors is interpatient heterogeneity^2^. Interpatient heterogeneity can be addressed by subtyping brain tumors to stratify patients and identify the best-matched drugs and therapies for a particular patient or cohort of patients. Molecular profiling analysis of a growing number of patients has revealed the full extent of GBM tumor heterogeneity. The genetic characteristics of GBM, such as the co-deletion of chromosomes 1p and 19q^7–9^, mutations in the genes coding tumor protein P53 (*TP53*)^10–12^, isocitrate dehydrogenase 1/2 (*IDH1/2*)^8,13^, epidermal growth factor receptor (*EGFR*)^14,15^, and deletion of the phosphatase and tensin homolog (*PTEN*)^14,16^ have refined the classification of GBM subtypes. Based on newer molecular and genetic data, the 2021 WHO classification of adult gliomas now defines GBM as any IDH wild-type diffuse glioma with presence of microvascular proliferation or necrosis and presence of *TERT* promoter mutations, or, gain of chromosome 7 and loss of chromosome 10 (i.e., 7+/10-), or, *EGFR* amplification. For sake of consistency with prior work especially the TCGA nomenclature, we have decided to use “GBM” with the caveat that some of the tumors in this cohort may not meet the criteria of how GBM is currently defined. To help the reader relate the findings we have provided a mapping of the new classification across all relevant results throughout the paper by using previous annotations^17^ (**Supplementary Table 9**). Analysis of gene expression and copy number alterations of GBM cataloged in The Cancer Genome Atlas (TCGA) has revealed several molecular subtypes: mesenchymal, proneural, and classical. Distinct clinical outcomes associated with these subtypes offer a stratification scheme to select appropriate therapies, albeit not yet at the resolution of N-of-1 personalized medicine. For instance, the proneural subtype is associated with a more favorable outcome, partly due to the high frequency of mutations in the *IDH1/2* genes. In contrast, the mesenchymal subtype is associated with worse survival than other subtypes^18–21^. Although *IDH* mutations have been linked to metabolic reprogramming, epigenetic reprogramming, and redox imbalance, the mechanisms by which *IDH* mutations predict favorable disease outcomes are not clearly understood^22^. Similarly, there is no integrated view of how diverse prognostic markers act in combination to influence clinical outcomes, including response to therapies, in an individual GBM patient.

Dysfunction across multiple pathways significantly shapes etiology and progression of GBM. Consequences of these mutations manifest through a network of interactions marked by non-linear dynamics, rendering the prediction of disease progression and therapy responsiveness challenging. To address the complexity of GBM and better characterize interpatient heterogeneity in GBM, our focus was on generating systems-level understanding of how dysfunction in diverse genes acts causally and mechanistically through a network of interactions to ultimately influence oncogenic processes and clinical outcomes. Previously, we developed SYstems Genetic Network Analysis (**SYGNAL**) to mine multi-omics and clinical outcome datasets for 422 GBM patients in TCGA to construct a causal and mechanistic transcriptional regulatory network (CM-TRN) for GBM (**gbmSYGNAL**)^23^. gbmSYGNAL delineated putative mechanisms by which 102 somatically mutated genes and pathways causally perturbed regulation by 74 transcription factors (TFs) and 39 miRNA regulators of approximately 5,000 genes across 500 disease-associated co-regulated gene modules (regulons). Through analysis of gbmSYGNAL, we uncovered novel insights, such as mechanism by which mutations in *NF1* and *PIK3CA* increase tumor lymphocyte infiltration by causally inducing the expression of *IRF1*, which upregulates a regulon of 27 genes associated with antigen processing and presentation. Furthermore, the network model generated actionable insights into the formulation of synergistic anti-proliferative and pro-apoptotic interventions with combinations of targeted inhibitors, anti-miRs, miRNA overexpression, and siRNAs against TFs^23^. Here, we report the construction of a revised TRN, called gbmMINER, which was generated using an updated version of SYGNAL that leverages the **M**echanistic **I**nference of **N**ode-**E**dge **R**elationships (**MINER**) algorithm^24^ to analyze multi-omics (whole exome sequencing, microarray, and RNA-seq) and clinical (cancer subtype and overall survival) data from a cohort of 516 patients. By outperforming cMonkey 2^25^ in accurately grouping 9,728 genes into 3,797 regulons, MINER significantly improved upon gbmSYGNAL by expanding the recall rate of TFs (from 6 to 26%) and miRNAs (from 15 to 25%) based on experimental support. Importantly, using a network quantizing technique to cluster regulons based on similar activity profiles, we identified 179 transcriptional programs that stratify GBM into 23 disease states. Using the quantized network, we developed prognostic models that were significantly more accurate than a previously best performing model based on gene expression correlates at predicting risk of disease progression. Notably, regulons and programs that significantly contributed to risk of disease progression provided hypotheses for mechanisms by which known prognostic markers influence clinical outcomes. Finally, through high-throughput (HTP) drug screening, we demonstrate proof-of-concept for prioritizing FDA-approved drugs based on activity profiles of regulons and programs containing relevant drug targets that accurately predicted, without any prior training, the drug sensitivity of 43 patient-derived glioma stem-like cells (PD-GSCs) to 28 anticancer drugs. Together the extensive validations demonstrate that gbmMINER represents an atlas of causal and mechanistic drivers of interpatient heterogeneity in GBM.

## RESULTS

### gbmMINER delineates how well known and novel disease driving mutations causally and mechanistically stratify GBM patients into 23 distinct states

We applied a new version of systems genetic network analysis (SYGNAL) that leveraged Mechanistic Inference of Node Edge Relationships (MINER) to generate a TRN by integrating multi-omics data and clinical outcomes from a cohort of 516 GBM patients in the TCGA^26^. In constructing gbmMINER, a total of 9,728 genes across the cohort were clustered by MINER into 3,797 modules of genes (henceforth “regulons”) with significant co-expression across subsets of patients and co-regulated by a common transcription factor or a miRNA (see Methods). The authenticity of regulons was ascertained by reproducing significant co-expression of member genes across at least one of three independent cohorts of 285^27^, 252^28^ and 80^29^ patients (**Supplementary Table 1**). Each regulon was quantized into three network activity states -- overactive, neutral, or underactive--based on statistical assessment of whether the expression levels of regulon member genes in a given patient were in the upper, middle, or lower thirds of the distribution of expression levels of each respective gene across all patients in the TCGA cohort. A notable advancement over the previous gbmSYGNAL model^23^, network quantization (see Methods) in gbmMINER maintained large-scale patterns of gene expression across the cohort, while significantly improving signal to noise^24^. In contrast to gbmSYGNAL, which identified 500 disease-relevant biclusters (avg. regulon size: 36 genes), gbmMINER identified 1,083 disease-relevant regulons (avg. regulon size = 11 genes) based on co-regulation of member genes in at least one independent dataset, and their association with patient survival (Cox HR p-value ≤ 0.05) or enrichment for a hallmark of cancer (enrichment p-value ≤ 0.05). (**Table 1**). To reduce the dimensionality of the TRN, the 3,797 regulons were clustered based on similar activity profiles across all patients into 179 distinct transcriptional programs, of which 58 programs contained the 1,083 disease-relevant regulons (**Figure 1A** and **Supplementary Table 1**). Similarly, based on correlated activity profiles of the 179 programs, the 516 patients were stratified into sub-populations of 23 distinct transcriptional states, potentially reflecting different subtypes of GBM disease-relevant expression signatures.

**Figure 1.**
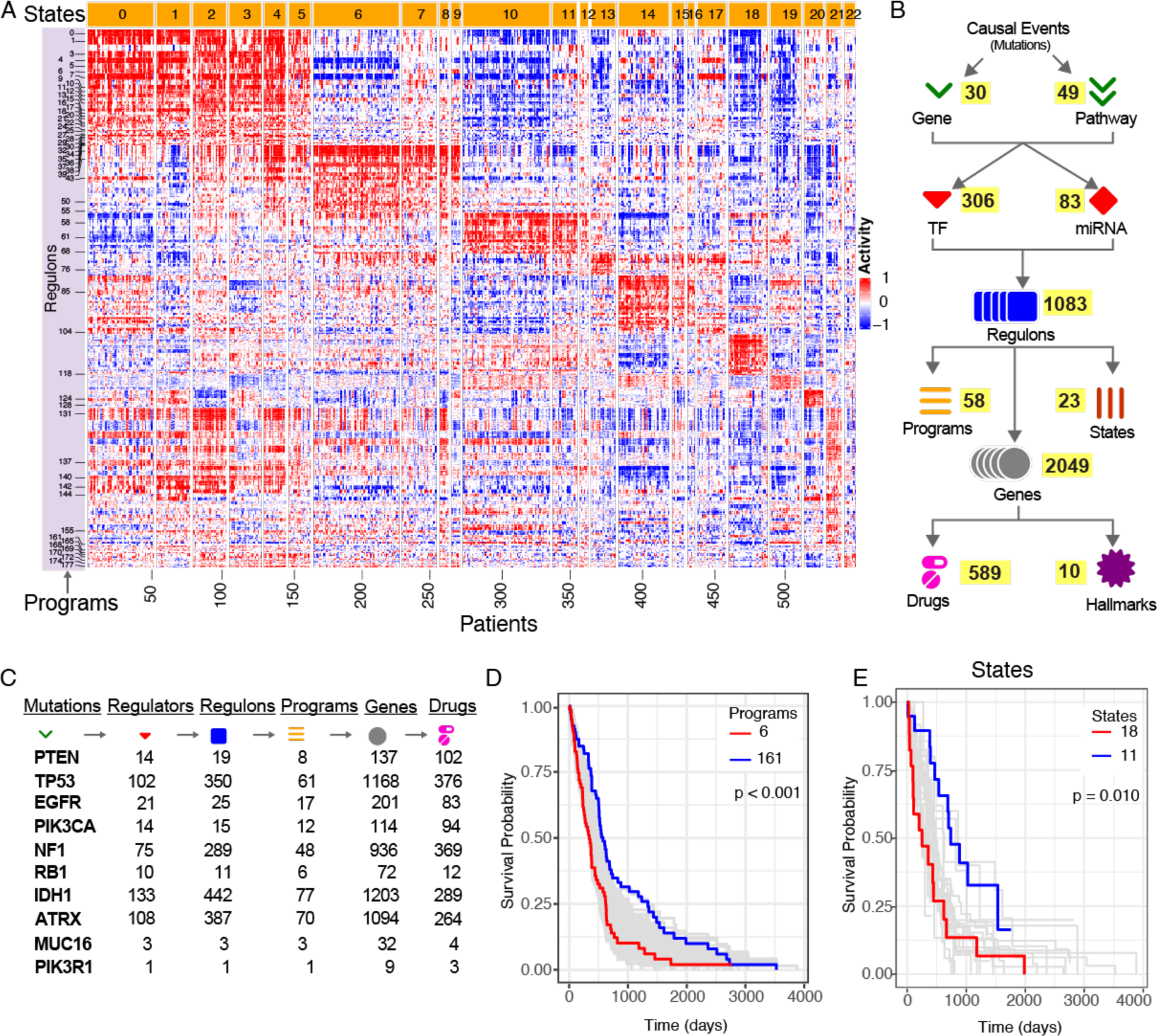
gbmMINER: a systems scale genotype to phenotype map of causal and mechanistic drivers of clinical outcomes in GBM. **A)**. Heatmap of regulon activity across patients reveals distinct transcriptional states wherein patients have very similar regulon activity across the entire network. Disease relevant programs, which are group of regulons with similar activity across patients, are indicated on the left while transcriptional states are shown on the top. Overactive, neutral and underactive regulons are colored in red, white and blue, respectively. **B).** The inferred gbmMINER TRN is a predictive map that implicates specific somatic mutations in causally modulating the expression of a TF(s) or miRNA(s) that in turn regulates genes within disease-relevant regulons. A summary of the counts for each feature in the gbmMINER TRN is shown. **C)** Causal and mechanistic relationships for known and novel GBM prognostic markers are well-represented in the model. **D)** gbmMINER transcriptional programs and **E)** states stratify risk of disease progression in TCGA cohort patients. Kaplan-Meier plot of survival probabilities for all programs and all states together with comparison of high-risk programs/states (red) versus low-risk programs/states (blue) are shown.

**Table 1.** Comparison of gbmSYGNAL and gbmMINER.

| Model Features | gbmSYGNAL | gbmMINER |
| --- | --- | --- |
| Total regulons | 1,830 | 3,797 |
| Average number of genes per regulon | 36 | 11 |
| Number of disease relevant regulons | 500 | 1,083 |
| Number of disease relevant programs | 58 | NA |
| Number of somatic gene/pathway mutations | 33/69 | 30/49 |
| Number of disease-relevant TF regulators | 74 | 306 |
| Number of disease-relevant genes | 5,193 | 2,049 |
| Number of miRNA regulators | 39 | 73 |
| TFs validated with CRISPR-Cas9 screening of patient-derived glioma stem-like cells<sup>31</sup> | 29<br>(p-value=0.24) | 137<br>(p-value=3.1e-5) |
| TFs validated with CRISPR-Cas9 screening of patient-derived glioma stem-like cells<sup>32</sup> | 26<br>(p-value=0.01) | 91<br>(p-value=0.002) |
| Overlap with GBM-relevant TFs in DisGeNET<sup>33</sup> | 16<br>(p-value=5.2e-4) | 114<br>(p-value=5.4e-21) |
| miRNAs with experimental-support for association with GBM (HMDD<sup>34</sup>) | 13<br>(p-value=2.6e-9) | 22<br>(p-value=6.2e-14) |
| Overlap with miRNAs dysregulated in GBM (miR2Disease<sup>35</sup>) | 7<br>(p-value=4.5e-7) | 7<br>(p-value=4.2e-5) |

Using the transcription factor binding site database (TFBS_db)^23^ and the Framework for Inference of Regulation by miRNAs (FIRM)^30^, 306 TFs and 83 miRNAs were implicated as likely regulators of the 1,083 disease-relevant regulons (**Table 1**, Methods). Further, somatic mutations within 30 genes and 49 pathways were causally associated with both the altered expression of regulators and concordant downstream consequences on mechanistic regulation of 2,049 genes within disease-relevant regulons associated with patient survival. Specifically, the causal influences of the mutations on disease-relevant regulons were confirmed to act through their mechanistic regulators by assessing differential expression of regulons across wild type and mutated patient samples, with and without perturbed expression of the regulators (**Figure 1B**, **Table 1** and **Supplementary Table 2**). The causal influences of 21 somatic gene mutations in gbmMINER were also previously modeled in gbmSYGNAL (overlap p-value:4.4e-32). Notably, gbmMINER modeled the causal influences of nine well-known driver mutations in GBM, including *EGFR*, *IDH1*, *NF1*, *PIK3CA*, *PIK3R1*, *PTEN*, *RB1*, *TP53*, and *ATRX* (**Figure 1C and Supplementary Table 5**) as well as 15 mutations with some known association with GBM. gbmMINER implicated causal and mechanistic diseases associations for 6 mutated genes (*DNAH2*, *APOB*, *TCHH*, *HSD17B7P2*, *KEL*, *KRTAP4-11*) that were not previously associated with GBM. Whereas most prognostic mutations for GBM were previously identified using GWAS, which does not provide mechanistic insights into their role in disease etiology or progression, gbmMINER delineated causal and mechanistic pathways through which mutations in these genes perturb the regulation of TFs and miRNAs, ultimately impacting the expression of downstream genes associated with the disease (**Figure 1C** and **gbmMINER Portal**).

### gbmMINER implicates novel transcription factors and miRNAs in GBM

Altogether, 306 TFs were implicated by gbmMINER in the regulation of disease-relevant regulons, which was a significant improvement over gbmSYGNAL, which had previously identified 74 TFs, of which 38 TFs were in both models (p-value = 7.4e-11). Phenotype data for 1,543 TF knockouts (94% of known TFs) from a genome-wide CRISPR-Cas9 screen^31^ identified 568 TFs had significantly altered proliferation in at least one of 10 patient-derived glioma stem-like cell lines (PD-GSCs). Of these 568 anti-proliferative TFs, only 29 TFs were in gbmSYGNAL (p = 0.24), whereas gbmMINER identified a total of 137 TFs (p = 3.1e-5), including 48 TFs that also altered the proliferation of human fetal neural stem cells (**Table 1** and **Supplementary Table 3**). Another CRISPR-Cas9 screen study on 2 PD-GSCs^32^ also validated 26 and 91 TFs identified in gbmSYGNAL (p=0.01) and gbmMINER (p=0.002), respectively. In addition, according to the DisGeNET database^33^ of disease-to-gene associations, 114 of the 306 TFs identified by gbmMINER (p = 5.4e-21), as compared to 16 of the 74 TFs identified in gbmSYGNAL (p = 5.2e-4), have important functions in GBM (See **Supplementary Table 3** for validation results). In summary, the gbmMINER network implicated 306 TFs in the regulation of 2,049 GBM-relevant genes, recapitulating 114 TFs that had been previously implicated in GBM, and 192 TFs with potentially novel and yet to be characterized roles in modulating disease outcomes in GBM.

gbmMINER also implicated 73 miRNAs as likely disease drivers based on their enriched binding sites in the 3’ UTRs of genes within disease-relevant regulons and their negatively correlated expression levels (R ≤ −0.2, p-value ≤0.05)^36^, which represents a significant improvement over the 39 miRNAs identified by gbmSYGNAL. Two lines of evidence supported the biological and disease relevance of miRNAs in the gbmMINER network. First, 22 of the 73 miRNAs (p-value=6.2e-14) were implicated in GBM in HMDD^34^, a curated database of evidence-based disease associations of human miRNAs (**Table 1**). Second, 7 miRNAs were implicated in GBM in the miR2Disease database^35^, which documents evidence for miRNAs that are dysregulated in human diseases (p-value=4.2e-5). In total, 25 of the 73 miRNAs had been previously implicated as dysregulated or causally associated with GBM, indicating the potential for discovering novel biology associated with the additional 48 miRNAs identified by gbmMINER.

### A network of master transcriptional regulators governs GBM transcriptional states, whose association with higher disease risk is correlated with increased immune evasion

Based on similarity of transcriptional program activities in gbmMINER, the 516 patients in the TCGA cohort were grouped into 23 transcriptional states. Notably, Kaplan-Meier survival analysis demonstrated that both programs and transcriptional states were associated with distinct overall survival outcomes (**Figure 1E** & **Figure 2B**). For transcriptional states we also mapped subtype information that was available for a significant number of tumor samples from 343 patients revealing that most of the astrocytomas were included in low to moderate risk states (**Figure 2A**). Using the ESTIMATE algorithm^37^, we calculated the ImmuneScore for each transcriptional state and investigated the relationship between immune cell infiltration and transcriptional states. The percentage of tumor-infiltrating immune cells correlated directly with the increased risk association across states (**Figure 2C**). Additionally, we tested each state for distinct mechanisms of tumor immune evasion by using the TIDE algorithm^38^. In general, higher risk states correlated with higher immune dysfunction in contrast to immune exclusion indicating that even though there was increased immune cell infiltration in higher risk states, those cells were in a dysfunctional state (**Supplementary Figure 9**). Based on estimation of the immune cell fractions from the RNASeq data^39^, there was no evidence of overrepresentation of any particular immune cells across states. No correlation was observed also between known prognostic markers, subtypes, and states, indicating that disease-associated pathway and gene mutations alone were not sufficient to determine the transcriptional state of a patient. For example, well known GBM prognostic markers such as *PTEN*, *TP53*, and *EGFR* were enriched across most states, while other markers were mainly enriched in states with a moderate risk association (data not shown). These findings suggested that the transcriptional states likely manifest from combinations of mutations acting through a complex network of regulatory interactions with system wide consequences on the activity levels of multiple disease-associated transcriptional programs.

**Figure 2.**
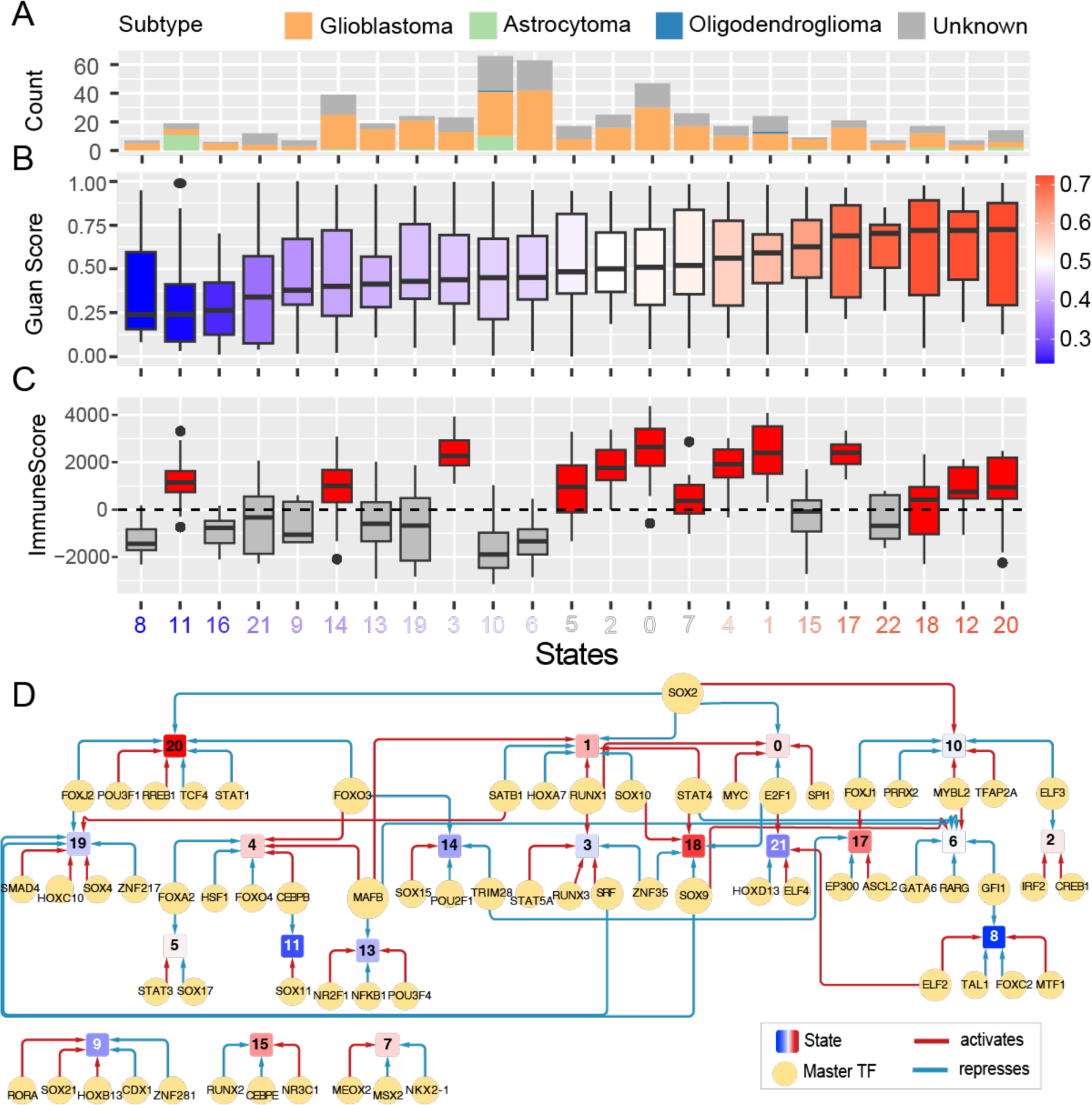
Association of disease risk of each transcriptional state with immune cell filtration and an elaborate network of master transcriptional regulators. **A)** Distribution of patients with different disease subtypes according to 2021 WHO classification ^17^ across each state. **B)** Boxplots of GuanRank risk scores for patients within transcriptional states, rank ordered from low to high median risk (from L to R, respectively). **C)** Boxplots of ImmuneScores indicating relative immune cell infiltration in tumors of patients within each state. **D)** Sixty-seven master TFs act in a combinatorial scheme to drive distinct transcriptional programs that characterize 20 of the 23 transcriptional states of GBM.

To better understand the transcriptional drivers of the 23 distinct states, we built a TF–TF network and shortlisted master TFs based on the ratio of outdegree/indegree edges (See Methods). We identified a network of 67 master regulators that were implicated in driving or suppressing each state (See Methods; **Figure 2D**). Most master regulators were specific to a given state, and only a few master regulators, including MAFB, FOX3, RUNX1, STAT4, SOX2, SOX9, E2F1, were shared across two or more states. Notably, knockdowns in 38 of 67 master regulators altered the proliferation of at least one PD-GSC (p= 1.4e-4) in a genome-wide CRISPR-Cas9 screen on 10 PD-GSCs^31^. In addition, 32 of the 67 master regulators (p = 5.4e-9) have important functions in GBM, such as in GSC self-renewal and maintenance (e.g., SOX2 and SOX9^40,41^ and MYC^42^, and repression of GBM cell differentiation (e.g., E2F1^43^, based on disease-to-gene associations in DisGeNET^33^. Despite extensive combinatorial control, including the influence of TFs that were uniquely associated as master regulators of some states, there was some concordance between type of influence (activator or repressor) of some master regulators on states with respect to their disease risk association.

### Program activities delineate known and novel biological processes underlying disease prognosis

Of the 179 programs in gbmMINER, Cox HR analysis implicated 58 as disease-associated, of which 7 programs were associated with low-risk (i.e., programs with negative hazard ratios that predicted good prognosis) and 51 programs were associated with high-risk (i.e., a positive hazard ratio) (**Figure 3A**, top panel). While the low-risk programs were enriched for oxidative phosphorylation (Pr-118), G2-M checkpoint (Pr-61), and Myc targets (Pr-144), the high-risk programs were enriched for hypoxia (15 programs, including Pr-172, Pr-6, Pr-43), epithelial-to-mesenchymal transition (20 programs including Pr-172, Pr-32, Pr-6), TNF-α signaling (28 programs including Pr-172, Pr-32, Pr-6), and other immune-related processes (**Supplementary Figures 1 and 3**). These results are consistent with significantly better survival of a newly discovered mitochondrial subtype of GBM that is characterized by oxidative phosphorylation^44^. In contrast, the mesenchymal subtype was associated with worse survival than other subtypes and was characterized by hypoxia and epithelial-mesenchymal transition^18–21^. The low-risk programs were significantly associated with GBM markers (*IDH1*, *ATRX*, *TP53* and *PDGFRA*) implicated in good prognosis, in contrast to high-risk programs that were enriched for mutations in *NF1*, a well-known negative prognostic marker for GBM (**Figure 3A** “**Mutations**” panel). Notably, relative to low-risk programs a significant number of high-risk programs also had higher ImmuneScores, which was consistent with the established association of increased immune cell infiltration and bad prognosis in GBM^45^ (**Figure 3A**, “**ImmuneScore**” panel). Notably, patients with overactive high-risk programs with high immune score also had a higher immune dysfunction score indicating that their bad prognosis was more likely associated with an impaired immune response. In contrast, we observed that patients with overactive low risk programs had lower immune dysfunction score and they had slightly increased CD4+ (non-regulatory) and CD8+ T-cells with decreased M1 and M2 macrophage levels (**Supplementary Figure 10**).

**Figure 3.**
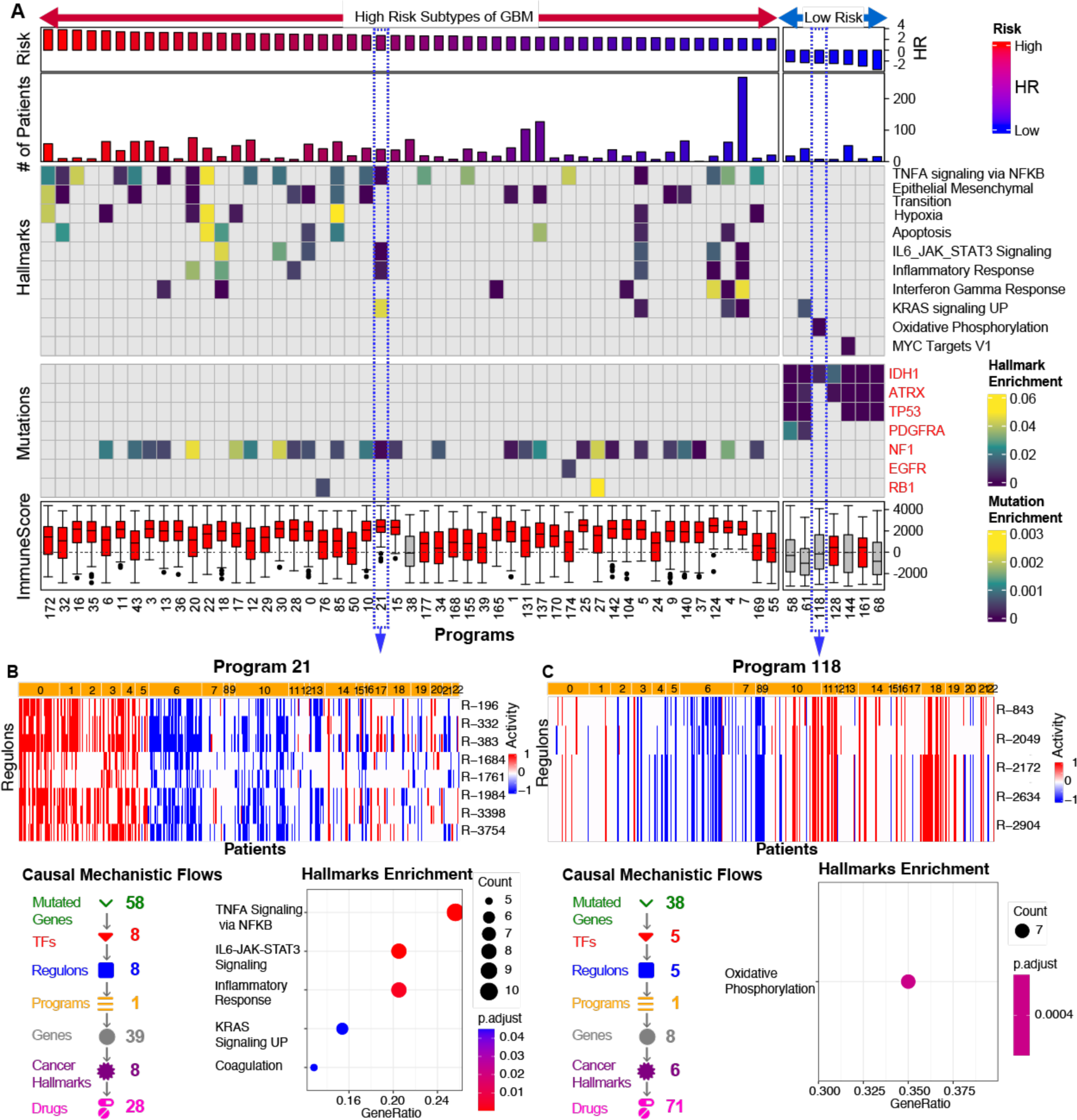
Casual and mechanistic underpinnings of high and low risk association of programs. **A)** Enrichment of disease hallmarks (upper heatmap) and mutations (lower heatmap) across disease associated programs. For each program, risk association and size (numbers of genes) are shown as barplots (top panels). Additionally, the level of immune cell infiltration for each program is indicated as a boxplot at the bottom (red: median ImmuneScore > 0, gray: median ImmuneScore <= 0). Detailed view of a high-risk programs 21 **(B)** and 118 **(C)**. The heatmap of the network activity of regulons within each program, with red for overactivity, white for neutral and blue for under activity (top panel). Summary of causal mechanistic flows associated with each program is shown in lower left panel. A dot plot of disease hallmark enrichments is shown in the lower right panel.

Through delineation of causal-mechanistic information flow (**CM-flow**) maps, gbmMINER uncovered insights into how specific mutations influence clinical outcomes by acting causally through TFs and miRNAs that mechanistically regulate activities of distinct biological processes. For example, 9 regulons within the high-risk program 21 (Pr-21; enriched for genes associated with TNF-α signaling, IL6-JAK-STAT3 signaling and inflammatory response), including regulons R-383, R-1984 and R-3754, were overactive across patients in most high-risk states.

gbmMINER predicted that mutations in NF1 causally activated TFs SATB1, ELF1, FOSL1, RUNX3 and ZNF232, which in turn upregulated 35 genes within regulons R-1684, R-332, R-1984, R-196 and R-1761 that were associated with TNF-α signaling, IL6-JAK-STAT3 signaling and inflammatory response, providing a causal and mechanistic hypothesis for the elevated ImmuneScore in these patient samples (**Figure 3B**). Conversely, the positive prognosis of *IDH1* mutations could be explained by their predicted causal inhibition of ETV7, which was implicated as an activator of Pr-118 regulons with genes of oxidative phosphorylation (**Figure 3C** and **Figure 5A**).

### Transcriptional program activities predict survival risk

Molecular markers such as *IDH1/2* mutations and *MGMT* promoter methylation sub-stratify patients with significantly better clinical outcomes and response to TMZ (*IDH* mutant vs WT: 31 months vs 15 months^22^, *MGMT* promoter methylation vs WT: 21.7 months vs 12.7 months^46^). However, 8% of *IDH1/2* WT patients in the TCGA cohort have had longer survival times (mOS: 44.6 months, range: 31-89 months) than patients with *IDH1/2* mutations and 23% of *MGMT* unmethylated patients survived longer (mOS: 42.7 months, range: 22-117 months) than *MGMT* methylated patients. Based on the observation that expression of individual regulons, transcriptional programs, and states were significantly associated with distinct survival outcomes, we explored if these features could serve as better prognostic markers of GBM (**Methods**). We used ridge regression to predict the risk of median overall survival with program activity (See **Methods**). First, we transformed patient survival data into GuanRank scores between 0 and 1^47^, with values > 0.5 indicating high-risk and values < 0.5 indicating low-risk. Using the concordance index (C-index) metric^48,48,49^ we evaluated the performance of each model on independent test datasets of 113 patients in TCGA dataset (20% of data not used to train the model) and 150 patients from an independent study (“Gravendeel dataset”^28^) and 80 patients from a nationwide observational study (“XCELSIOR study”, see Methods). We also compared the gbmMINER risk prediction models to the performance of a 4-gene-panel model that stratifies patients based on a risk score calculated using the sum of weighted expression levels of four autophagy genes: *DIRAS3, LGALS8, MAPK8,* and *STAM*^49^. The performance of the gene panel in risk stratifying patients was significantly lower with C-index values of 0.58, 0.57 and 0.56 for the TCGA, Gravendeel and XCELSIOR datasets, respectively (**Figure 4A-B** and **Table 2, Supplementary Table 8**). Notably, the KM plot for TCGA and XCELSIOR patients stratified by the gene panel was not statistically significant. By contrast, performance of the program model was consistently better across all datasets. The predicted low- and high-risk classes by the program model also effectively stratified patients based on their survival outcomes, as shown by significant separations in the survival curves of these individual risk groups in the KM plot (**Figure 4A-B)**. Furthermore, we assessed performance of the program risk model relative to risk stratification by *IDH* mutations and *MGMT* promoter methylation status within the XCELSIOR cohort. As expected, *MGMT* promoter methylation status stratified *IDH* Wild type patients into low and high-risk groups **(Figure 4C)**. Remarkably, the gbmMINER program risk model also effectively stratified patients into low and high-risk groups, regardless of their *IDH* mutation and *MGMT* promoter methylation status **(Figure 4D)**. Importantly, the program risk model also sub-stratified *MGMT* methylated patients, identifying low risk patients who were likely super-responders to standard of care, and high risk patients who may benefit from combining standard of care with 2^nd^ line treatments **(Figure 4E)**.

**Figure 4.**
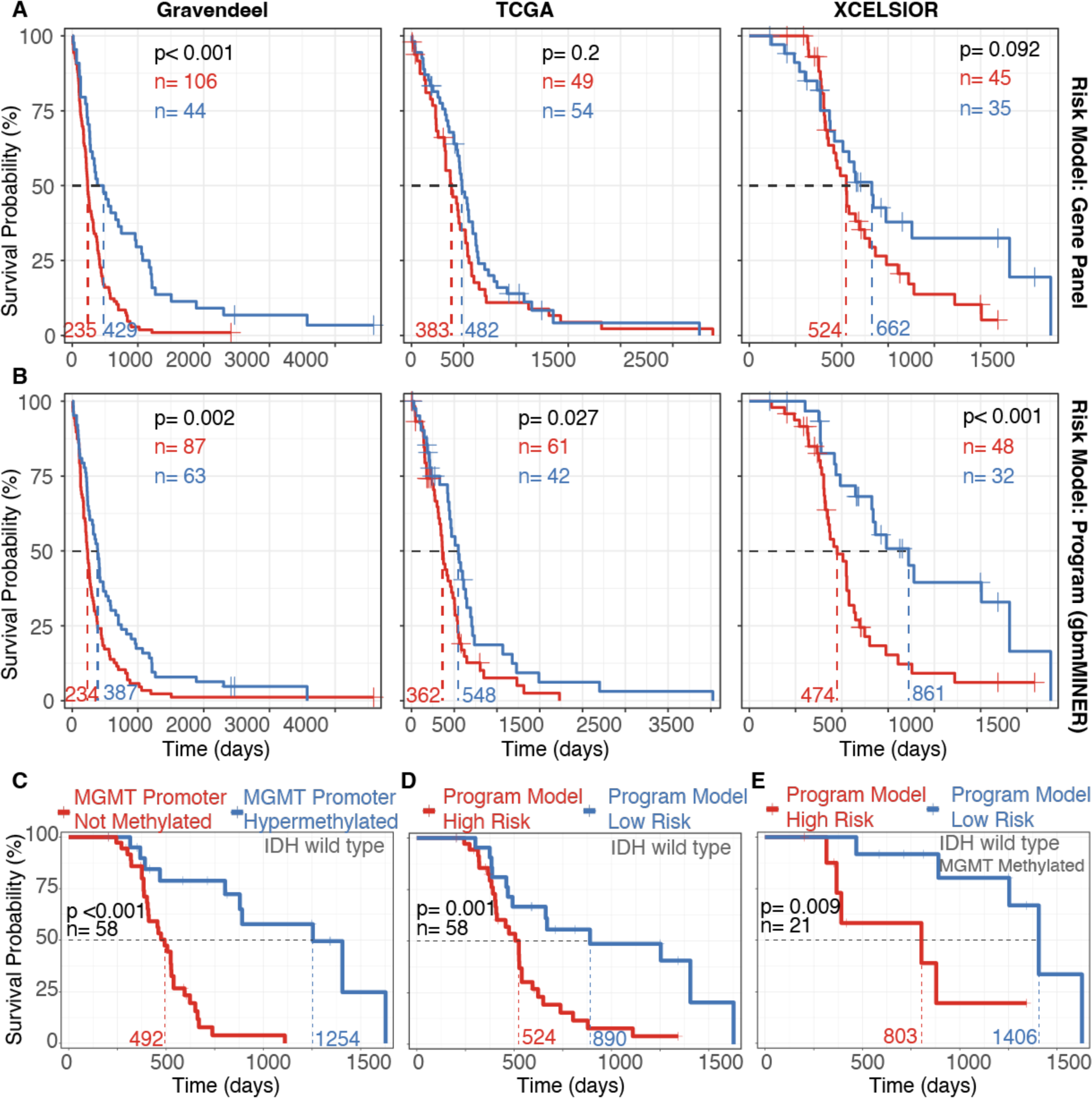
gbmMINER-based risk prediction models outperform previously best performing gene panel and performs as good as IDH WT and MGMT Promoter Methylation based stratification. The stratification of predicted low-risk (blue) and high-risk (red) groups by Kaplan-Meier curves for TCGA, Gravendeel, and XCELSIOR datasets across **A)** A gene panel-based risk prediction model ^49^ (yellow panel header background and **B)** The gbmMINER based program risk prediction model (blue panel header background). For each plot C-index, number of patients, mOS and log-rank test p-values for the significance of the survival probabilities between the high and risk groups are shown. (See **Supplementary Table 8** for risk model outputs and survival analysis results). Stratification of *IDH* Wild Type XCELSIOR cohort patients by C) *MGMT* Promoter Methylation status and **D)** gbmMINER program risk model. **E)** Sub-stratification of *IDH* Wild type and *MGMT* Promoter Hypermethylated patients by program risk model.

### Better prognosis of *IDH1*, *ATRX* and *TP53* mutations likely manifests from modulation of ferroptosis through their causal influences on ETV7 and CTCF regulons

gbmMINER uncovered many of the well-known positive and negative prognostic markers for GBM that act through a combinatorial scheme to modulate risk-associated programs. For instance, the network model revealed a complex combinatorial scheme in which 15 mutations causally perturb the expression of 12 TFs that mechanistically co-regulate 34 genes (**Figure 5**). Notably, this CM-flow map captured how well-known mutations, including *IDH1*, *TP53*, and *ATRX*, might causally influence clinical outcomes through seven TFs (ETV7, POU3F2, POU3F3, CTCF, HOXA3, MEF2A, and NR3C1) that combinatorially regulate eight genes. Of the seven regulators, POU3F2, a neurodevelopmental TF^41,50^, and HOXA3, an activator of aerobic glycolysis, have been implicated in promoting GBM propagation^51^. The expression of these seven TFs was predicted to be causally influenced by nine somatically mutated genes, of which mutations in *IDH1* (p-value:2e-10), *ATRX* (p-value:5e-6), and *TP53* (p-value:0.006) were associated with Pr-118 overexpression (**Figure 5A**).

**Figure 5.**
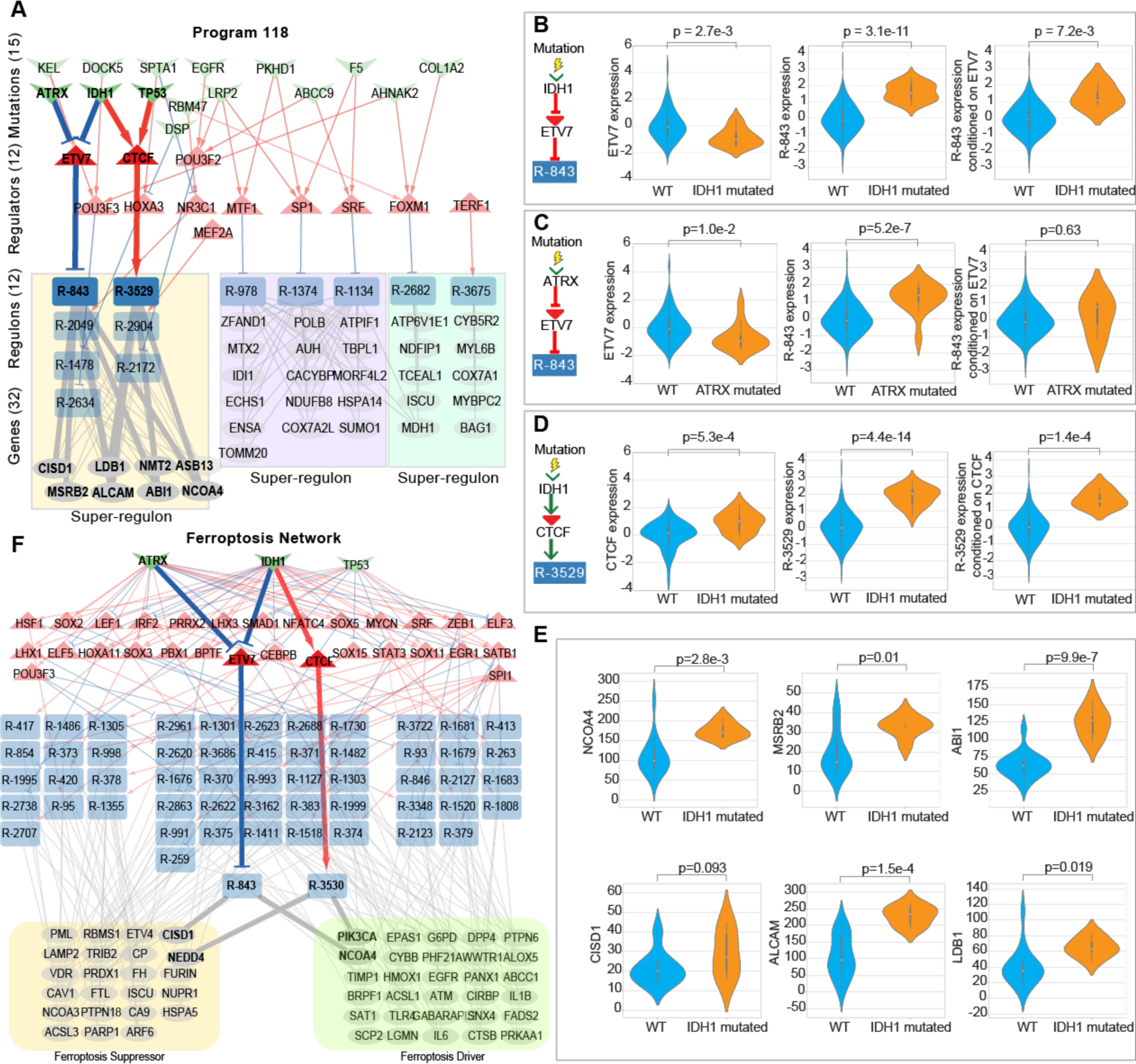
*IDH1*, *ATRX* and *TP53* mutations causally impact ETV7 and CTCF to modulate ferroptosis leading to better prognosis. **A)** Network diagram of causal and mechanistic influences of mutations and TFs on Program 118. Regulons comprising program 118 are represented as blue boxes with their associated regulators (red triangles) and putative causal genetic abnormalities (green chevrons). Red edges denote activation and blue edges denote inhibition. Regulons with significant overlap of member genes are grouped into super-regulons (yellow, purple and green rectangles). **B)** ETV7 mediates the causal influence of somatically mutated *IDH1* on downstream genes in regulon R-843. ETV7 expression (left boxplot) and expression of genes in R-843 (middle boxplot) increases when *IDH1* is mutated. Conditioning R-843 expression on ETV7 abolishes the increase in expression indicating that the causal influence of IDH1 is mediated by ETV7. **C)** ETV7 mediates the causal influence of somatically mutated *ATRX* on downstream genes in regulon R-843. **D)** CTCF mediates the causal influence of somatically mutated *IDH1* on downstream genes in regulon R-3529. **E)** ETV7 regulon genes are upregulated in *IDH1* mutants. **F)** Combinatorial scheme through which *IDH1*, *ATRX*, and *TP53* mutations modulate 29 TFs implicated in mechanistically regulating 53 ferroptosis genes. The 23 ferroptosis driver genes are grouped within the yellow shaded box and 30 ferroptosis suppressor genes in the green shaded box.

From the CM flows, we extracted the novel insight that *IDH1* and *ATRX* mutations repress ETV7, thereby activating a super-regulon of eight genes within Pr-118 (**Figure 5A-C)**. Note that by design MINER reconstructs multiple regulatory influences on the same set of co-regulated genes by discovering redundant instances of the same regulon, each associated with a unique regulatory influence. Combining these redundant regulons gives a super-regulon, which is so called because it is implicated to be under the control of a large number of regulators-in this example, 7 TFs were assigned as regulators of the super-regulon of 8 genes.

Across significant numbers of patients in the TCGA dataset, the expression of these 7 TFs was inferred to be causally perturbed by at least 10 somatically mutated genes. While not previously implicated in GBM, ETV7 has been shown to regulate breast cancer stem-like cell features by repressing IFN-response genes^52^ and activating mTORC3 assembly to promote tumor growth in a mouse model^53^. In addition, the model predicted that *IDH1* and *TP53* mutations act through CTCF to activate the same super-regulon (**Figure 5A, C** and **D and Supplementary Figure 4**). This finding explains why loss of CTCF binding sites, which modulates communication between enhancers and promoters, is associated with *IDH* and *TP53* mutated GBM tumors^54^. Within this super-regulon, two of the eight genes, *NCOA4* and *CISD1*, are related to ferroptosis (enrichment p-value = 0.002)^55–58^. Specifically, whereas NCOA4 increases free iron levels in cells and promotes ferroptosis, CISD1 reduces mitochondrial iron uptake^57^. A third super-regulon gene, *MSRB2*, encodes methionine-R-sulfoxide reductase a multifunctional enzyme that both scavenges reactive oxygen species and is involved in the biosynthesis of cysteine, a key component of GSH, which regulates ferroptosis^57^. All genes in the ETV7 regulon were significantly upregulated in *IDH1* mutants (**Figure 5E**), a key finding that was also reproduced in an independent cohort^59^ (**Supplementary Figure 5**). The mechanistic hypotheses were validated by the CRISPR-Cas9 screen^31^, which showed that ETV7 significantly altered proliferation in one glioblastoma stem cell, POU3F2 altered proliferation in three glioblastoma stem cells, and CTCF altered proliferation in seven glioblastoma stem cell lines. A number of mechanisms have been proposed for the improved prognosis of patients with *IDH1/2* mutated GBM^22^. Our findings are consistent with these prior explanations and extend our mechanistic understanding of how *IDH1/2* mutations may causally inhibit ETV7 to activate ferroptosis and improve prognosis (i.e., median survival of ∼31 months as compared to ∼15 months for *IDH* wildtype GBM^22^).

Further, the combinatorial influences of *IDH1*, *TP53* and *ATRX* mutations on regulation of ferroptosis offered a plausible explanation for why *IDH1* mutant gliomas are genetically associated with *TP53* mutations or *ATRX* mutations^60–62^. To investigate these influences on a systems level, we mined gbmMINER for the entire causal-mechanistic network linking the three mutations to ferroptosis-related TFs that were implicated in regulating at least two genes implicated as drivers or suppressors of ferroptosis as per FerrDb database^63^. We also required ferroptosis-related TFs to have significantly altered proliferation of at least one PD-GSC in the CRISPR-Cas9 screen^31^. Based on these criteria, *IDH1*, *TP53* and *ATRX* mutations were implicated in causally influencing the expression of 29 ferroptosis-related TFs of which 14 were previously implicated in regulating ferroptosis (**Supplementary Table 4**).

The 29 TFs were implicated in mechanistically regulating 53 ferroptosis-related genes across 18 super-regulons. 23 of the 53 genes were implicated as suppressors of ferroptosis and the remainder were implicated as ferroptosis drivers (**Figure 5F**). For example, a known ferroptosis pathway, HSF1-HSPB1, increases the resistance of cancer cells to ferroptosis through the inhibition of iron uptake^58^. In addition to the 14 known regulators of ferroptosis, gbmMINER predicted that this process was also putatively regulated by an additional 15 TFs, including ETV7, HOXA11, LEF1, LHX1, LHX3, NFATC4, BPTF, POU3F3, ELF3, SOX11, CTCF, SOX3, SOX5, SRF, and ELF5 (**Supplementary Table 4**). Consistent with their gbmMINER predicted roles in activating ferroptosis-driver genes, ELF5, SOX11 and HOXA11 were previously demonstrated to act as tumor suppressors in many cancers^64–68^. Conversely, consistent with its gbmMINER-predicted suppressor role, NFATC4 is known to promote oncogenesis^69^. Finally, the integrated analysis demonstrated that mutations in *IDH1* (11 ferroptosis-activating and 5 ferroptosis-suppressing CM-flows), *ATRX* (8 ferroptosis-activating and 7 ferroptosis-repressing CM-flows) and *TP53* (8 ferroptosis-activating and 5 ferroptosis-suppressing CM-flows) were the major modulators of ferroptosis in GBM. Because *IDH* mutations can fundamentally change the biology of the disease, and given the low numbers of *IDH* mutant glioma patients in our cohort, we sought to ascertain generalizability of our findings to a larger cohort of 514 low grade glioma (LGG) patients by analyzing multiomics data from the TCGA-LGG cohort and constructing a lggMINER model. Analysis of the lggMINER model independently confirmed that IDH1 mutations do indeed causally influence regulation of ferroptosis genes in LGG patient tumors by modulating a similar set of TFs (**Supplementary Figure 12**). In summary, gbmMINER delineated how mutations in *IDH1*, *ATRX* and *TP53* causally and mechanistically modulate specific cancer related processes, including ferroptosis, to have prognostic implications on clinical outcomes.

### Network quantization and drug-constrained regulon activity uncovers patient-specific disease-network map to enable therapy prioritization

A major hurdle in uncovering predictive biomarkers of drug response for personalized medicine applications is the insufficiency of HTP screening datasets for most drugs across a diversity of patients, patient-derived xenografts or tumor cells, with paired pre- and post-treatment genomic/transcriptomic profiles. Using patient derived glioma stem-like cells (PD-GSCs), we investigated if activity of transcriptional programs (or regulons) containing drug target(s) would accurately predict drug sensitivity of a patient’s tumor cells. In brief, RNA-Seq profiles of 43 PD-GSCs were analyzed through network quantization with gbmMINER to generate PD-GSC-specific disease network maps. In brief, the entire disease-perturbed network within a PD-GSC was uncovered by treating each regulon as a discrete unit and evaluating whether it was overactive, underactive, or neutral (using a p-value cutoff of 0.05) based on the distribution of z-scored expression levels of member genes. Drug constrained regulon activity (DCRA) for a given drug was then calculated as the mean activity of all regulons containing or regulated by its target(s), and compared to the background distribution of DCRA values for that drug across all patient samples in a cohort (e.g., TCGA cohort). In doing so, we used DCRA to estimate the overall status of the disease-relevant networks targeted by a given drug and thereby predict whether a given PD-GSC was likely to be a responder (low IC_50_) or non-responder (high IC_50_) to treatment with that drug. Specifically, if DCRA was greater than zero for an antagonist drug (less than or equal to zero for an agonist drug), then the PD-GSC was predicted to be sensitive to that drug. By contrast, if the reverse was true then the PD-GSC was predicted to be a non-responder. Using this approach, we predicted sensitivity of 43 PD-GSCs to 62 drugs based on patterns of over- and under-activity of regulons containing cognate drug targets (Methods). Notably, this analysis revealed that the distributions of DCRAs for most drugs across the PD-GSCs was significantly skewed relative to corresponding distributions in the TCGA cohort (**Supplementary Fig 6**). This skewed distribution could be attributable to inherent increased or decreased drug susceptibility of PD-GSCs or the absence of tumor microenvironment influences. Nonetheless, the predicted susceptibilities of 43 PD-GSCs to 62 drugs was then compared to responder/non-responder classifications based on IC_50_ values that were experimentally determined in a HTP drug screen (**Methods** and **Figure 6A**).

**Figure 6.**
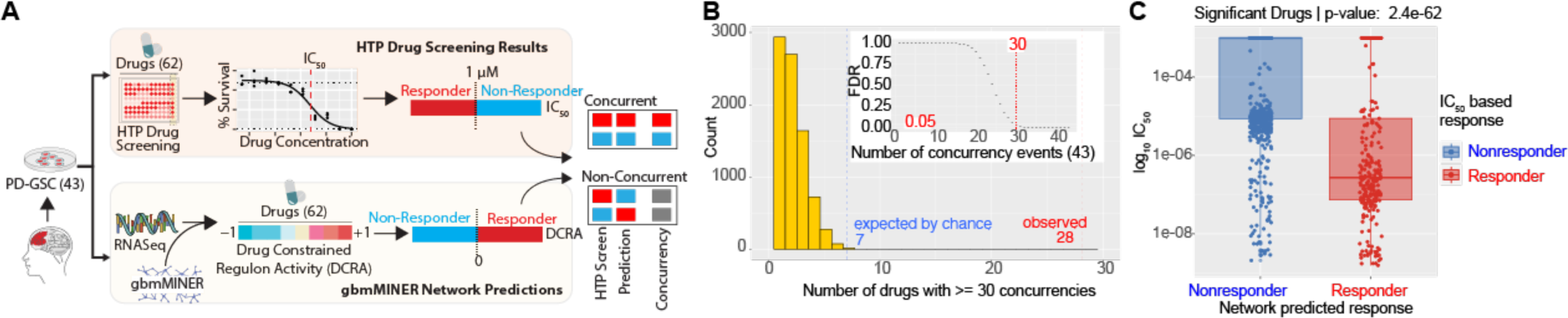
Network quantization uncovers patient-specific disease networks enabling N-of-1 prediction of therapy responsiveness. **A)** Schematic illustration of the workflow for validating network-based therapy response predictions through concordance analysis against PD-GSC high throughput drug screening results. **B)** Statistical significance of concurrency was determined through a permutation test. The inset plot shows distribution of the single drug FDR adjusted p-values up to 43 concurrencies. At least 30 concurrencies are needed (red dashed lines) to achieve FDR < 0.05. The histogram in the main plot shows that by chance a maximum of seven drugs would be expected to have >= 30 PD-GSCs that have concurrent predicted and experimentally-measured sensitivities (green dashed lines), while 28 (red dashed lines) drugs were observed to have >=30 concurrencies in the actual comparison (p-value = 0e-1 as determined with permutation testing). **C)** Comparison of the experimentally-determined IC50 values for predicted responders and nonresponders for significantly concordant drugs. p-values were determined with a Mann-Whitney U test.

While a significant number of drugs had variable efficacy across PD-GSCs, many drugs had consistently high (i.e., low IC_50_) or low (i.e., high IC_50_) efficacy across most of the 43 PD-GSCs, which was consistent with their skewed distributions of DCRA values relative to the TCGA cohort. Importantly, the predicted sensitivity of the PD-GSCs were significantly concordant with the experimentally determined IC_50_ values. Based on permutation tests, it was determined that at least 30 concordances between predicted and experimentally determined response to each drug would be necessary to be considered statistically significant alignment at an FDR < 0.05. For a dataset of 43 PD-GSCs against 62 drugs, while we would expect by chance that just 7 drugs would show ≥30 concordances with experimental results, we observed efficacy predictions for 28 drugs agreed with experimentally determined sensitivity of up to 43 PD-GSCs (p-value=0, **Figure 6B, Supplementary Figure 8**). The distinct DCRA-predicted sensitivity profiles across these drugs for each PD-GSC was consistent with the HTP screen, which demonstrated significant difference in IC_50_ values between drugs that were predicted to be effective or not-effective for each PD-GSC (**Figure 6 C**).

Given that the MTT assay estimates cell viability, we investigated whether the concordance of PD-GSCs vis-à-vis predicted and measured drug sensitivity was explained by the activity status of drug-target containing regulons enriched for genes associated with the viability-relevant cancer hallmark “evading apoptosis”. Indeed, responder PD-GSCs that were concordant (i.e., with high DCRA and low IC_50_ values) had a high proportion of drug target containing regulons (59%) that were overactive and enriched for genes associated with “evading apoptosis”. The converse was also true, that is non-responder PD-GSCs that were concordant (i.e., low DCRA and high IC_50_ values) were associated with a greater proportion (61%) of drug-target containing regulons that were underactive and enriched for “evading apoptosis” genes (**Supplementary Figure 11**). These results suggest that drugs which induce cytotoxic effects have higher concordance between DCRA and MTT viability-assay-based assessment of sensitivity, and that discordant drugs may target other functions that do not necessarily lead to cell death. If so, then the accuracy of drug sensitivity predictions by gbmMINER could turn out to be even higher if IC_50_ of discordant drugs are estimated using biochemical assays that probe relevant biological functions enriched in regulons containing their respective target(s).

## DISCUSSION

We have generated a model for characterizing interpatient heterogeneity in GBM. The model was demonstrated to have high mechanistic accuracy based on extensive performance evaluation of independent patient cohorts, disease databases, literature, CRISPR-cas9 phenotypic screens from two independent studies on 12 PD-GSCs and normal neuronal cell lines, and through HTP screening of 62 drugs against 43 PD-GSCs. Findings from these studies demonstrated the remarkable mechanistic accuracy with which gbmMINER has mapped modular architecture of gene-gene co-regulation within regulons, identifying TFs and miRNAs that differentially regulate cancer processes across sub-groups of GBM patients. In so doing, gbmMINER delineated networks through which well-known and novel somatic mutations in genes and pathways influence clinical outcomes and response to therapy. For instance, gbmMINER delineated how *IDH1*, *ATRX* and *TP53* mutations act through a network of 29 regulators to modulate 53 ferroptosis genes across three super-regulons, affecting treatment response and prognosis. In this manner, we can now interpret how 30 driver mutations modulate expression of 306 TF and 83 miRNA regulators to alter activity patterns of 179 transcriptional programs, 58 of which were determined to be disease-relevant. In so doing, gbmMINER has stratified GBM patients into 23 states with varied disease risk, which is mechanistically governed by a distinct TRN.

A prognostic model developed using TRN status (i.e., activity patterns of programs) was significantly more accurate and robust at predicting clinical outcomes (mOS), particularly in comparison to a 4-gene panel^70^. The higher performance of the transcriptional program-based risk model is attributable to the network quantization approach, which likely overcomes issues of variability in gene expression that may manifest from multiple sources, including technical issues such as sample processing and variabilities across profiling technologies, and biological issues such as disease stage of sampling, and composition of cells in patient sample. Specifically, since the program model averages expression values of co-regulated genes within modules, it likely overcomes noise and missing values in expression patterns for individual genes. Furthermore, the program model also provided insights into the biology of disease-driving mechanisms that distinguish low-risk patients from high-risk patients, regardless of their *IDH* mutation and *MGMT* methylation status. For instance, we discovered that *IDH1*, *TP53* and *ATRX* mutations act combinatorially to modulate up to 29 TFs and activate ferroptosis, which explains the better prognosis of *IDH* mutants (31 months of mOS compared to 15 months mOS for *IDH* WT)^22^.

Three principal mechanisms have been put forth to explain why *IDH* mutations predict favorable disease outcomes^22^. First, *IDH* mutations drive metabolic reprogramming by increasing oxidative metabolism and suppressing glutamine metabolism^22^. Indeed, we observed genes of oxidative phosphorylation were significantly enriched in program 118 modulated by *IDH1* mutations (p-value= 8.1e-07). Second, *IDH* mutations epigenetically reprogram tumor cells by inhibiting DNA and histone demethylation, leading to hypermethylation globally or at specific targets^22^. Consistent with this hypothesis, HM27 methylation profiling revealed that 3 ferroptosis-related TFs (LHX3, ETV7, PPRX2) were significantly hypermethylated at their promoters in *IDH1* mutant GBM^71^ (**Supplementary Figure 7**). At least in the case of ETV7, the hypermethylation of this TF was consistent with the repression of its expression levels in *IDH1* mutated GBM patients. Third, *IDH* mutations activate ferroptosis through accumulation of excessive reactive oxygen species^22^. Our findings integrate and extend the three mechanisms into a unified view that explains how *IDH* mutations perturb a network of TFs to modulate oxidative phosphorylation and DNA methylation, resulting in activation of ferroptosis.

We inferred CM-flows in the ferroptosis network from just 14 *IDH*-mutated tumor samples, within the TCGA-GBM cohort, that should be reclassified as low-grade gliomas. We reproduced these findings across risk-stratified patient subgroups in the 514 LGG patient dataset, including the causal influence of *IDH1* mutation on ETV7 and its downstream regulons. In addition to showcasing the sensitivity of gbmMINER in delineating disease-driving mechanisms in a small subgroup of patients within a larger cohort, this finding also underscores generalizability of the mechanisms by which *IDH1* acts through a combinatorial network to activate ferroptosis. While there is evidence in literature that both IDH1 and TP53 regulate ferroptosis, the primary mechanism was unknown^72^. In fact, a recent study discovered that although the acetylation-defective mutant TP53^3KR^ lost its ability to induce cell senescence, apoptosis, and cell-cycle arrest, it was still able to inhibit tumorigenesis by inducing ferroptosis ^73^. Activating the ferroptosis pathway is being explored as a promising therapeutic strategy for many cancers, including GBM, as cancer cells tend to have higher metabolic demand for iron^74^. The regulatory network of ferroptosis delineated in gbmMINER could facilitate such efforts. We demonstrated that quantized DCRA values of regulons and programs in gbmMINER accurately predict drug sensitivity based on the distribution of DCRA values across a reference cohort, without the need for machine learning on a large training dataset. This approach is also generalizable towards repurposing FDA-approved drugs with targets within disease-relevant regulons and programs in gbmMINER, for matching treatment regimen to a patient based on their DCRA profiles, and for identifying and prioritizing new drug targets.

## Data Availability

All data produced in the present study are available upon reasonable request to the authors

https://gbm.systemsbiology.net

## ACKNOWLEDGEMENTS

S.T. is supported by R01-CA259469, Washington Research Foundation Grant No: GA-00117 and Andy Hill Cancer Research Endowment (CARE) Fund. W.W., K.K. and C.M. are supported by R01-CA259469. M.M. is supported by R01-CA259469 and Washington Research Foundation Grant No: GA-00117. Y.H. is supported by R01-CA259469 and Andy Hill Cancer Research Endowment (CARE) Fund. A.N. is supported by NSF-2150265. A.P.P. is supported by R01-NS119650, the Burroughs Wellcome Career Award for Medical Scientists, and Discovery Grant from the Kuni Foundation. P.H. and H.L. were funded by the Ben and Catherine Ivy Foundation and are currently supported by R01-CA259469 and philanthropic funding from the Swedish Medical Center Foundation. C.C. is supported by R01-CA259469. J.P. was funded by a fellowship from the NIH (F32-CA247445) and currently supported by NIH grant R01-CA259469. N.S.B. is supported by R01-CA259469, Washington Research Foundation Grant No: GA-00117 and Andy Hill Cancer Research Endowment (CARE) Fund.

We thank Timothy J. Martins and the UW Quellos High-throughput Screening Core for advice and services in running the HTP screens.

## SUPPLEMENTARY INFORMATION

### Supplementary Tables

**Supplementary Table 1:** Summary of disease relevant regulons and validations across independent datasets

**Supplementary Table 2:** List and details of disease relevant causal mechanistic flows

**Supplementary Table 3:** Validation for the GBM association of 306 TFs.

**Supplementary Table 4:** TFs were revealed to be associated with ferroptosis in published studies

**Supplementary Table 5:** Known and new prognostic markers identified in gbmMINER

**Supplementary Table 6:** FDR values for a drug showing n (count) number of concurrent instances.

**Supplementary Table 7:** p-values for observing n (count) number of drugs showing at least 30 concurrent instances.

**Supplementary Table 8:** Risk prediction model and Survival analysis output

**Supplementary Table 9:** Clinical information about patient outcomes and tumor subtypes for TCGA cohort patients

### Supplementary Figures

**Supplementary Figure 1:** Enriched molecular hallmarks for significant risk-associated programs.

**Supplementary Figure 2:** Enrichment of genetic programs for Hallmarks of Cancer.

**Supplementary Figure 3:** KM stratification curves for Program 118.

**Supplementary Figure 4:** An insight that TP53 mutations activate CTCF and activate ferroptosis is extracted from program 118.

**Supplementary Figure 5:** Data from Trautwein et al., supports that ETV7 regulon genes are upregulated in IDH1 mutants.

**Supplementary Figure 6:** Comparison of PDGSC and TCGA training cohort drug constrained network activity (DCRA).

**Supplementary Figure 7.** LHX3, ETV7 and PPRX2 have significantly higher methylation at their promoters in IDH1 mutants.

**Supplementary Figure 8.** Distribution of the concurrencies for all the drugs tested in PD-GSC high throughput screen.

**Supplementary Figure 9.** Distribution of Immune Dysfunction scores and immune cells across states.

**Supplementary Figure 10.** Distribution of Immune Dysfunction scores and immune cells across programs

**Supplementary Figure 11.** Activity status of “evading apoptosis”-enriched regulons containing targets of drugs that were concordant between DCRA-predicted and MTT-assay determined drug sensitivity across relevant responder and non-responder PD-GSCs. “Concurrency percent” refers to the proportion of concurrency events between predicted and observed drug sensitivity.

**Supplementary Figure 12.** Overlap of gbmMINER and lggMINER network for ferroptosis related IDH mutation causal mechanistic flows.

## METHODS

### Glioblastoma somatic mutations, miRNA and mRNA expression data

Training data for GBM-MINER network construction was acquired from The Cancer Genome Atlas (TCGA). Gene expression microarray data were normalized and Z-scored. Gene expression RNA-seq data were TMM-normalized and Z-scored. A total of 536 patient tumors and four post-mortem controls containing gene expression data for 10,263 genes combined with both RNASeq and microarray Z-scored data were used to train the MINER model (**Supplementary Table 9**). The miRNA expression data for 536 miRNA samples were quantified by microarray and used for causal and mechanistic inference of miRNA regulation of co-expressed gene clusters identified by MINER as described before^23^. A gene was considered to be mutated in a patient’s tumor if a somatic mutation was observed that modified the gene coding sequence (missense, nonsense, frameshift, in-frame insertion or deletion, splice site, modified translation start site, introduced new start site, or removed stop codon). An NCI-nature pathway was considered to be mutated in a patient’s tumor if at least one gene member of the pathway had a nonsynonymous somatic mutation (all TCGA mutations were not annotated as silent). Mutated genes and pathways used in causal network inference were required to be mutated in at least 5% of the patient tumors.

### Glioblastoma clinical data for survival analyses

Glioblastoma clinical phenotypes for survival analyses were acquired from The Cancer Genome Atlas (TCGA) using Broad Firehose. Clinical information about patient outcomes and tumor subtype was used in Regulon post-processing (**Supplementary Table 9**).

### Independent glioblastoma validation gene expression cohorts

Raw Affymetrix HGU133 Plus 2.0 CEL files were either downloaded from the NCBI Gene Expression Omnibus (GEO; GSE7696 and GSE16011)^28,29^ or EMBL-EBI ArrayExpress (E-MTAB-3073)^27^. The CEL files for each of the three studies used in our analysis were background subtracted and quantile normalized using the justRMA function from the affy package of Bioconductor (www.bioconductor.org) using an alternative CDF file^75^. The 18,023 non redundant probes were selected by collapsing highly correlated probes (correlation coefficient ≥ 0.8) that mapped to the same Entrez ID as they are likely measuring the same transcript isoform. Only grade II, III and IV gliomas were included in the analyses. In addition to these three microarray datasets, RNA-seq data generated as part of the Ivy Glioblastoma Atlas Project (https://glioblastoma.alleninstitute.org/static/home) was TMM-normalized and used as an RNA-seq expression-based validation cohort. All validation data were Z-scored after normalization.

### MINER disease relevant regulon membership prediction validation

Regulons were considered significantly coexpressed if the variance explained by first principal component within coherent members was greater than or equal to 0.3 and was significantly larger than random samples (empirical p-value ≤ 0.05 and replication in at least one independent dataset empirical p-value ≤ 0.05; Each of the 3,764 regulons were post-processed to discover: (i) coexpression quality via variance explained by first principal component within coherent members (empirical p-value < 0.05 and variance explained ≥ 0.3), (ii) validation of co-expression quality in at least one independent GBM cohort (empirical p-value ≤ 0.05); (iii) putative TF or miRNA (via the FIRM pipeline) regulators associated with regulon with a minimum spearman’s correlation of 0.3 between regulators and eigen gene within coherent members (only negative correlation between miRNA and regulon) in the training dataset; (iv) survival analysis with regulon eigen genes (cox hazards ratio (HR) p-value ≤ 0.05); (v) independent validation of survival association in one of the three microarray datasets (cox HR p-value ≤ 0.05); (vi) association with hallmarks of cancer (Jiang-Conrath Semantic Similarity Score ≥ 0.8)^30,76^; and (vii) putative somatic gene mutations and pathways with T-test p-value <= 0.05 for regulators and regulon eigen gene between mutated and wild-type samples. The regulons were filtered using the above-mentioned criteria for validation of co-expression, mechanistic inference, causal inference and disease relevance (**Supplementary Table 1**). Recall rate of TFs between gbmSYGNAL and gbmMINER was calculated as the number of TFs supported by CRISPR and DisGeNet divided by total number of TFs. The recall rate for miRNA was calculated as the number of miRNAs supported by HMDD divided by the total number of miRNAs.

### Network quantization: Calculation of discrete regulon activity

Calculation of discrete regulon activity was performed as described before^24^. Briefly, for each standardized patient sample, gene expressions were organized from the least to the most expressed and divided into three equal segments: lower, middle, and upper thirds. Assuming effective normalization of gene expression, we can propose a null hypothesis suggesting that a random assortment of genes lacking any co-expression relationship will distribute evenly across these thirds. To assess this, we employ a binomial distribution with a probability parameter (p) of 1/3 to model the likelihood of ‘k’ genes falling within the same third, given a selection of ‘N’ genes, where ‘N’ is greater than or equal to ‘k’. A standard p-value of 0.05 is applied as a threshold for rejecting the null hypothesis, indicating that the selected gene set lacks co-expression. Genes surpassing this threshold in the lower third are designated as “under-active,” while those exceeding it in the upper third are labeled “over-active.” Instances falling outside these categories are denoted as “neutral.” Consequently, we generate a matrix assigning values of {-1, 0, 1} to represent the discrete activity of all regulons across all samples. This methodology for calculating discrete regulon activity has previously been employed in the MINER analysis for patients diagnosed with multiple myeloma ^24^, with adaptations made accordingly.

### Regulon clustering to identify super-regulons

For the 12 regulons in Program 118 and the 55 ferroptosis-related regulons, genes in each regulon were first converted into 1 (appearing) or 0 (non-appearing) using bag of words encoding method^77^. Then k-means clustering was used to cluster the encoded regulons. The optimal number of clusters (super-regulons) is determined by the combination of Elbow method and Silhouette method^78^.

### Risk prediction

We used ridge regression models to predict risk scores. TCGA dataset was randomly split into a training set (70%), validation set (10%), and test set (20%). The models were learned from the training set and evaluated on the test set and two independent datasets. The validation set was used to select the optimal hyperparameter in the ridge regression models. Based on three types of input data, gene expression (TPM), regulon activity (−1,0,1), and program activity (−1,0,1), we learned three different models and referenced them as gene, regulon, and program models, respectively. An extra parameter related to the number of features (genes/regulons/programs) was learned from the validation set, similar to the hyperparameter in ridge regression. The models output GuanScore, where a score <= 0.5 is predicted to be low risk, and a score > 0.5 is predicted to be high risk. Models were evaluated using the concordance index as well as Kaplan-Meier curves to estimate patient survival for the predicted risk groups in all datasets. The 4-gene-panel model used for comparison calculates the risk score using the sum of weighted expression levels of four autophagy genes [(0.1052 × [DIRAS3]) + (0.2152 × [LGALS8]) + (−0.3603 × [MAPK8]) + (−0.2851 × [STAM])].

### Identification of master regulators from TF-TF network

For each state, we first identify a list of TFs causally associated with the state and then build TF– TF network in three steps. First, Least Absolute Shrinkage and Selection Operator (LASSO) regression models are generated for each TF in the list to predict its expression using a subset of the other TFs in the list as predictors. Specifically, the TF list is a subset to include only TFs with a binding site for the target TF in TFBSDB or a CHiP-seq database. The second step is to prune the LASSO models to minimize the number of TF predictors necessary to maintain the same level of predictive accuracy. Finally, the LASSO coefficients of each predictor TF for each target TF are defined as weighted edges to connect the TFs in a network. After building TF-TF network, we rank all TFs by the ratio of outdegree/indegree and pick the top TFs as master regulators, because master regulator is defined as the gene at the top of the regulatory hierarchy, which should not be affected by the regulation of any other genes. A script for the whole process is provided.

### Drug mapping pipeline

OpenTargets (with 6515 drugs in the database) and the Cancer Cell Line Encyclopedia (CCLE) (with 24 drugs as part of drug screening) were the two main sources used to map therapies to our updated GBM network. Drugs were mapped to genes belonging to all levels of causal-mechanistic flows, namely, genes containing somatic mutations, transcription factors associated with regulons, and individual regulon genes, if the gene is targeted by the drug.

### Patient samples, cultivation of PD-GSCs, and high throughput drug screening

WHO grade IV glioblastoma tumors were obtained from surgeries performed at Swedish Medical Center (Seattle, WA) according to institutional guidelines. The freshly resected tumor tissues were processed to generate PD-GSCs as described^79^. For the 43 PD-GSCs used in this study, 39 were established as neurosphere cultures and maintained in ultra-low attachment dishes (Corning). The other 4 PD-GSCs grew as adherent monolayers in T75 flasks pre-treated with laminin (1:100; Sigma). All 43 PD-GSCs were cultured in serum-free media consisting of Neurobasal Medium-A (Gibco^TM^) with 2.0% (v/v) B-27 serum-free supplement minus vitamin A (Gibco^TM^), 20 ng/mL EGF (PeproTech Inc.), 20 ng/mL FGF-2 (PeproTech Inc.), 20 ng/mL insulin (Sigma), 1 mM sodium pyruvate (Corning), 2 mM L-glutamine (Gibco^TM^) and 1% Antibiotic-Antimycotic (Gibco^TM^). PD-GSC cultures were maintained at 37°C, 5% CO2, 1% O2, with culture pH monitored with phenol red. Cultures were refed every 2-3 days. High throughput (HTP) screening assays (IC_50_ studies) were conducted essentially as described^79^. In brief, PD-GSCs were added to either laminin coated 384-well plates (adherent monolayer cultures) or polyhema coated 384-well plates (neurospheres) at a density of ∼2000 cells per well using a Thermo Scientific Matrix WellMate. Following incubation of PD-GSCs overnight at 37 °C, drugs were added to individual wells using the CyBi-Well vario liquid handler (Analytik Jena) and plates were incubated at 37°C for 96 hours. Drugs were added at concentrations ranging from 0.0005 – 10 μM to generate 10-point dose response curves for each PD-GSC culture (n=43) and each candidate drug (n=62). Following the incubation period, CellTiter-Glo (Promega) was added to individual wells per the manufacturer’s recommendations and luminescence was measured on a Perkin Elmer EnVision plate reader. HTP measurements were collected in duplicate or triplicate (depending on PD-GSC cell counts), and data was corrected for background luminescence. PD-GSC cultures tested were within 6 passages from the initial GSC enrichment from the original tumor biopsy.

### Determination of IC_50_ values for each drug-PD-GSC combination

The IC_50_ values used in our analysis were generated by applying the ‘drm’ (“drc” R package^80^) log-logistic 4 parameter model (LL4) to the dose-response curve data (dose values and associated percent viability across 3 replicates) where all 4 parameters were estimated. We then estimated the IC_50_ values manually based on the LL4 model produced by the ‘drm’ function. The percent viabilities across the curve’s dose range were calculated by inputting values that covered the drug screen dose range into the LL4 regression model which then returned the associated percent viabilities at each point in the estimated dose response curve. The IC_50_ value was then identified by finding the dosage that returned a percent viability closest to the 50% viability value. The 50% viability value used to identify the IC_50_ dose value was calculated as half the mean percent viability of the 3 replicates at the lowest dose range.

### Calculating the concurrency between the gbmSYGNAL predicted and experimentally determined drug sensitivities of PD-GSCs

We first classified each PD-GSC/drug combo as to whether they responded to a drug (responder or nonresponder) for the GBM drug screen, and the gbmMINER drug sensitivity prediction. The IC_50_ was used to classify the PD-GSC’s in the drug screen while the drug constrained network activity was used to predict drug sensitivity (DCRA) for gbmMINER. Drugs are mapped to gbmMINER by identifying all regulons that either contain, or are regulated by, at least one of a drug’s targets and those regulons are then considered to be mapped to that drug. The gbmMINER DCRA for each PD-GSC/drug combination is calculated as the mean of all drug-mapped regulons and will range from −1 to 1. If both the GBM drug screen and the gbmMINER predictions show the same response (responder or nonresponder) then they are concurrent between the two measurements.

The statistical significance of the number of concurrency instances for each drug (x/43) and the entire drug screen was generated with permutation testing described in the following steps.

*Single drug FDR adjusted p-values for 1 to 43 number of concurrencies*.

1. Permute the classification labels (responder, nonresponder) for both the drug screen and the gbmMINER predictions. This creates a null distribution of responses based on the original results.
2. Calculate the concurrencies of the permuted null distribution in the same manner as with the original data for each drug/PD-GSC combination.
3. Repeat the permutation 10000 times.
4. For each drug (62 drugs) calculate the probability of seeing at least n (n=1 to 43) instances of concurrency across the 43 PD-GSCs exposed to that drug.

a. Sum number of times each drug showed at least n concurrent instances and divide by 10000.
b. This produces a probability for the number of concurrencies ranging from 1 to 43 for each drug and can be interpreted as a p-value. This created a 62 by 43 matrix where the rows are the 62 drugs, and the columns are the number of potential number of concurrencies out of the 43 PD-GSCs. Each cell has the p-value associated with the number of concurrencies of greater than or equal than the nth number of concurrencies.
c. FDR was applied across each column (62 drugs) to account for multiple testing.
5. The FDRs were highly consistent across all drugs for all n number of concurrent instances so the mean of the FDRs across the 62 drugs were used as the final FDR for each number of concurrencies (**Supplementary Table 6**).
6. A drug showing at least 30 (from **Supplementary Table 6**) instances of concurrency passed an FDR cutoff of 0.05.

*Significance of entire drug screen*

1. We found 28 drugs that showed at least 30 instances of concurrency in our original analysis.
2. We calculated the number of times we found a drug showing at least 30 instances of concurrency in each iteration of our permutation process and divided by 10,000.

a. This gives us a probability, interpretable as a p-value, that shows how likely we are to see at least n (1 to 62) number of drugs that showed at least 30 instances of concurrency (**Supplementary Table 7**).
3. A literal p-value of 0 was generated for the likelihood of seeing at least 28 drugs showing 30 instances of concurrency. In fact, the highest number of drugs that showed at least 30 instances of agreement out of 10000 iterations was 7 out of 62 drugs.

### N-of-1 therapy mapping for a patient

Causal mechanistic (CM) flows associated with disease-relevant regulons are shortlisted from all CM flows inferred by the gbmMINER. Therapy candidates are also shortlisted as those used as anti-cancer drugs in the past and/or used in GBM trials in the past according to clinicaltrials.gov. Regulon activity is calculated for a particular patient to show which regulons are over-or under-active among the disease-relevant regulons. Their associated CM flows are also shortlisted. Short-listed drugs were mapped to patient-specific CM flows. If a disease-relevant regulon is associated with a positive RMST difference (RMST higher in under-active patients compared to over-active patients), and if the regulon is over-active, as well as if the patient shows a mutation that upregulates regulon activity, shutting down the regulon genes might inhibit the cancer-causing pathway associated with the regulon genes. Hence, if a regulon gene is a target of a shortlisted drug that is an inhibitor of the gene, and if the gene is highly expressed, the drug is a good therapeutic candidate for the patient and may be used for further testing. Similar conclusions of potential agonists or antagonists can be drawn with different combinations for RMST differences, regulon activity, and mutations affecting regulon activity. Similar inferences have been made for the TF regulator target of a drug. In the case of mutations, if mutations present in a patient cause gene amplification and the mutation is causally associated with upregulation of a regulon that shows poor survival in upregulated patients, an inhibitor acting against the mutated gene might be a promising target.

### XCELSIOR Observational Study

The XCELSIOR real-world registry is a pan-cancer observational study (<u>NCT03793088</u>) that started in 2019. Patients may enroll via electronic consent and sign a blanket HIPAA release to permit aggregation of electronic medical records and structured EMR data from all sites of care. Unstructured text from clinic narratives were utilized as source documents for annotation in an electronic database. Raw genomics and transcriptomics data (FASTQ files) were collected from commercial labs. Dates of diagnosis were abstracted from pathology reports. Patient death dates were verified for accurate overall survival calculations. For patients without known death date, censoring was based on the most recent of the document date or last medication record available in EMR. In the current study, clinical and omics data from 80 patients were used.

---

**Supplementary Figure 1.**
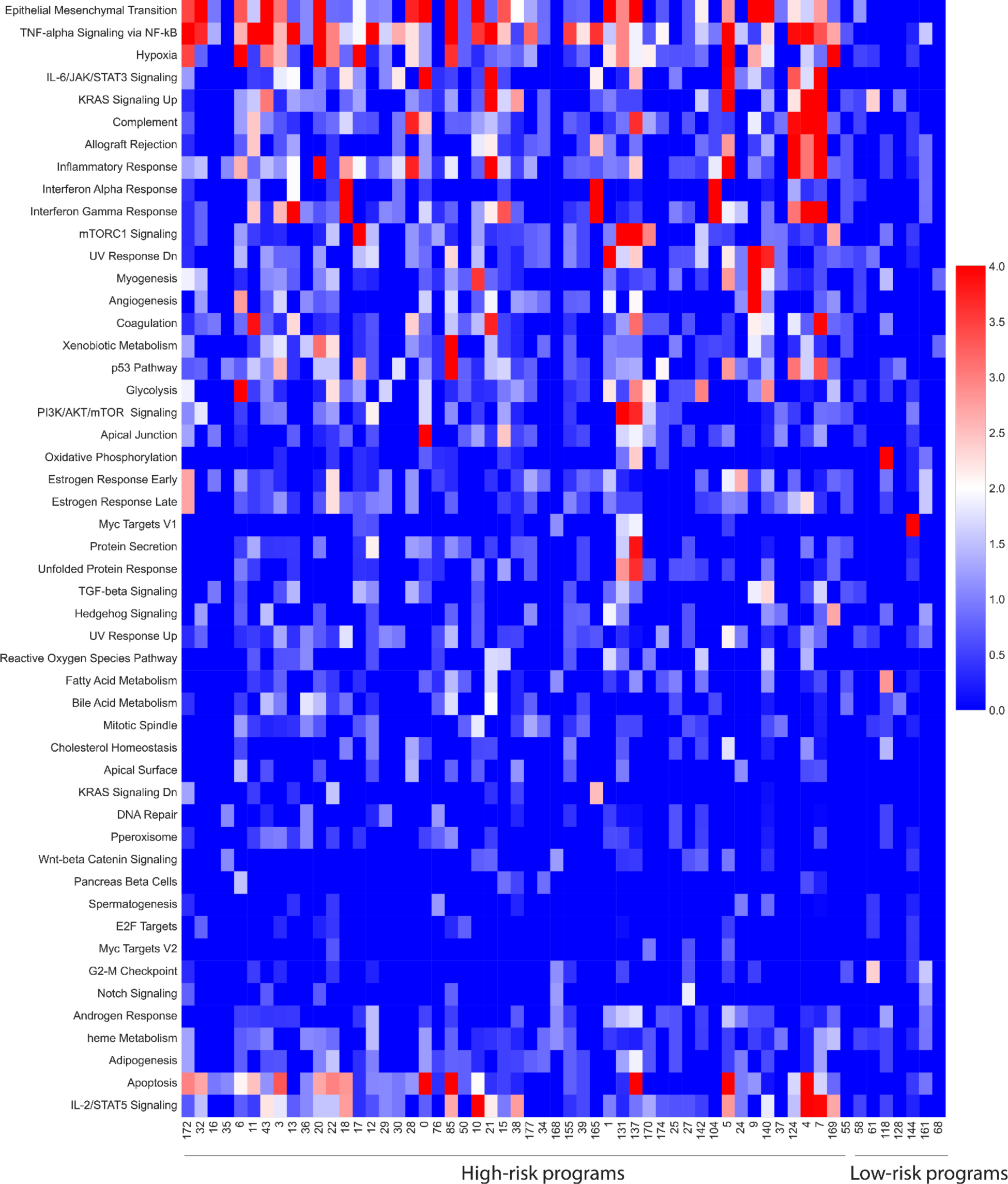
Enriched molecular hallmarks for significant risk-associated programs. Low-risk programs are mainly enriched for Oxidative Phosphorylation and Myc Targets. High-risk programs are mainly enriched for hypoxia, EMT, TNF-alpha Signaling, and immune responses.

**Supplementary Figure 2.**
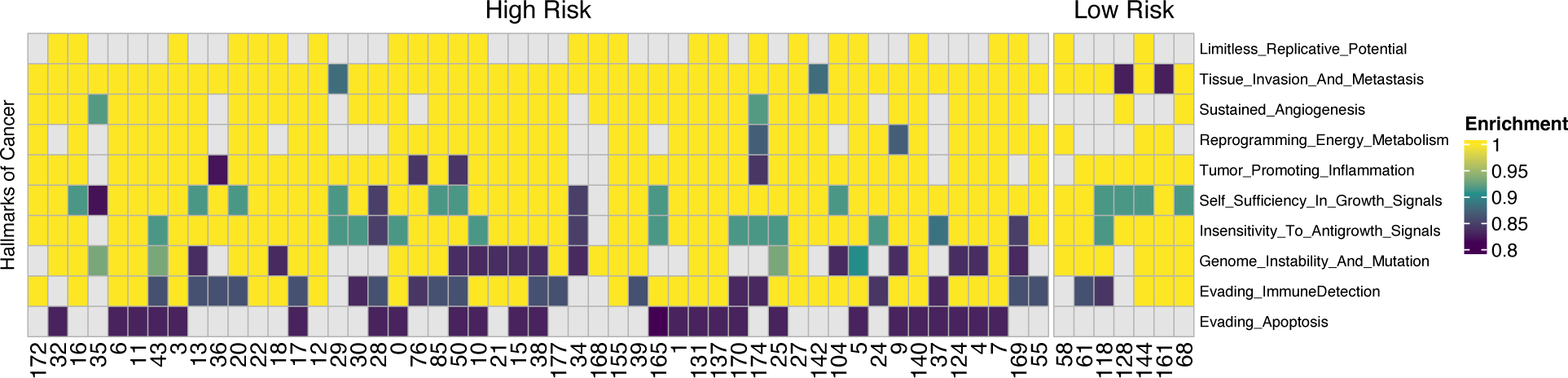
Enrichment of genetic programs for Hallmarks of Cancer. Enrichment for each program was calculated by using semantic similarity of the GO terms associated with genes in the programs.

**Supplementary Figure 3.**
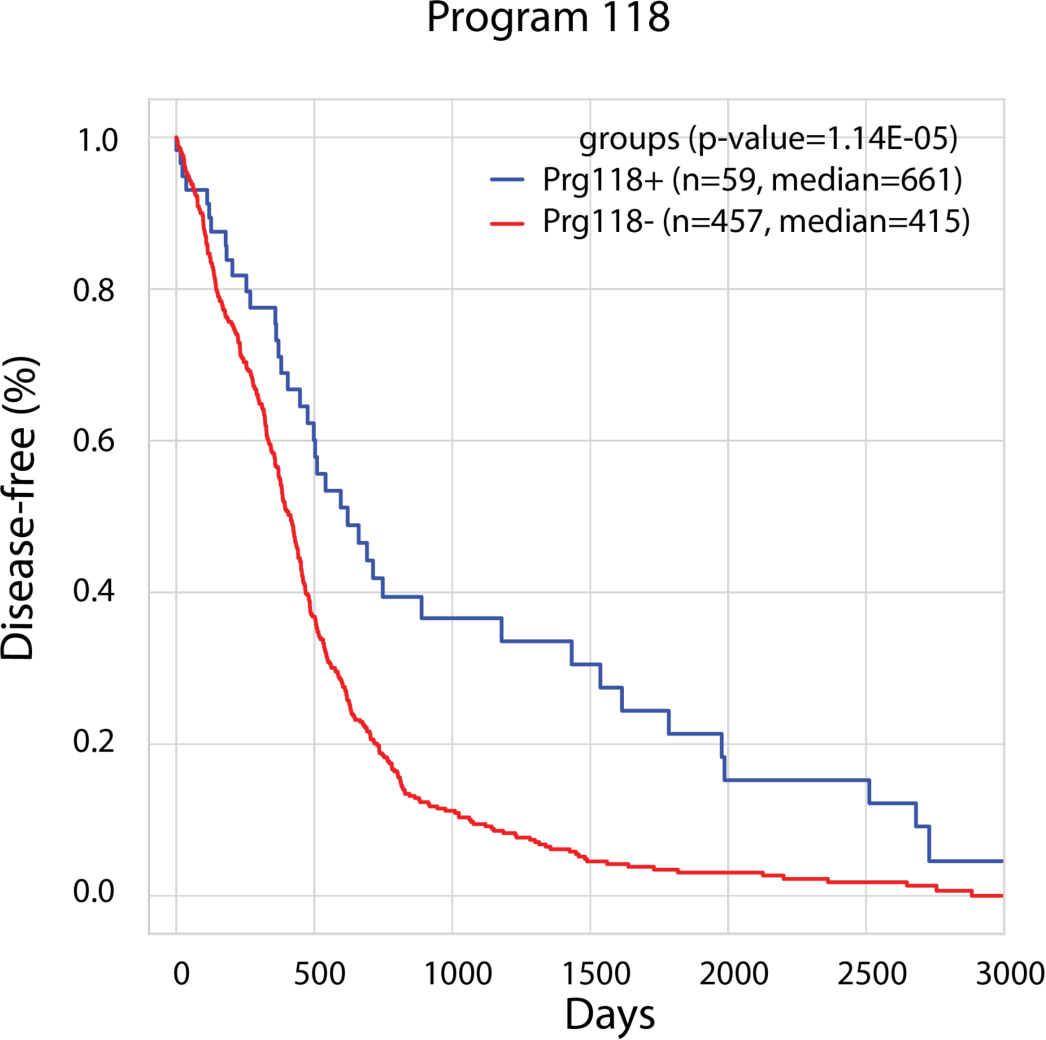
KM stratification curves for Program 118. Program 118 is a top program that stratifies GBM patients in low-risk programs.

**Supplementary Figure 4.**
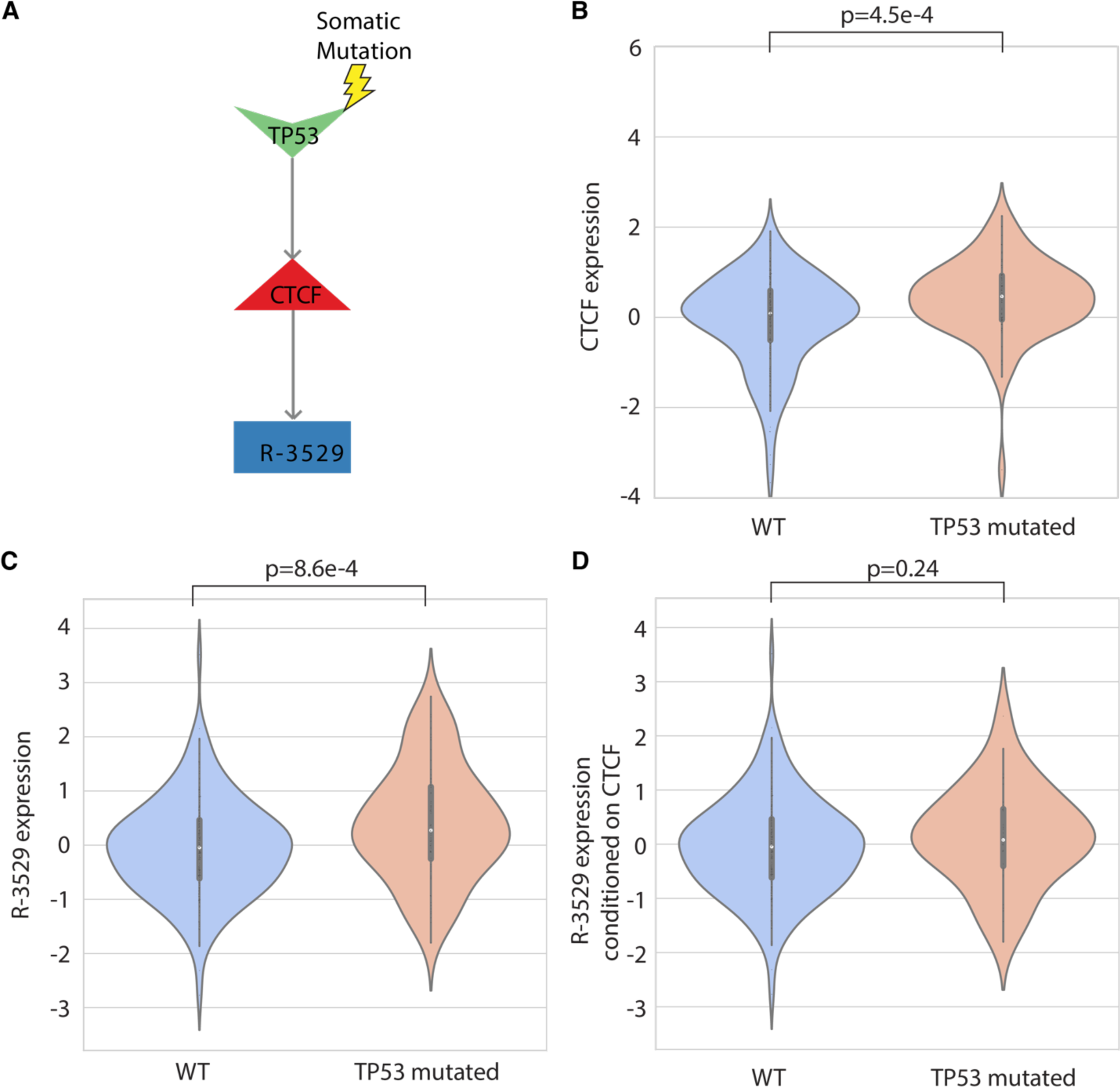
An insight that TP53 mutations activate CTCF and activate ferroptosis is extracted from program 118. **A)** Extracted insight for TP53 mutations from program 118. **B)** CTCF expression is significantly upregulated when TP53 is somatically mutated. **C)** The expression of CTCF regulon is significantly upregulated when TP53 is somatically mutated. **D)** Conditioning CTCF regulon expression on the expression of CTCF destroys the link between CTCF regulon and mutations in TP53.

**Supplementary Figure 5.**
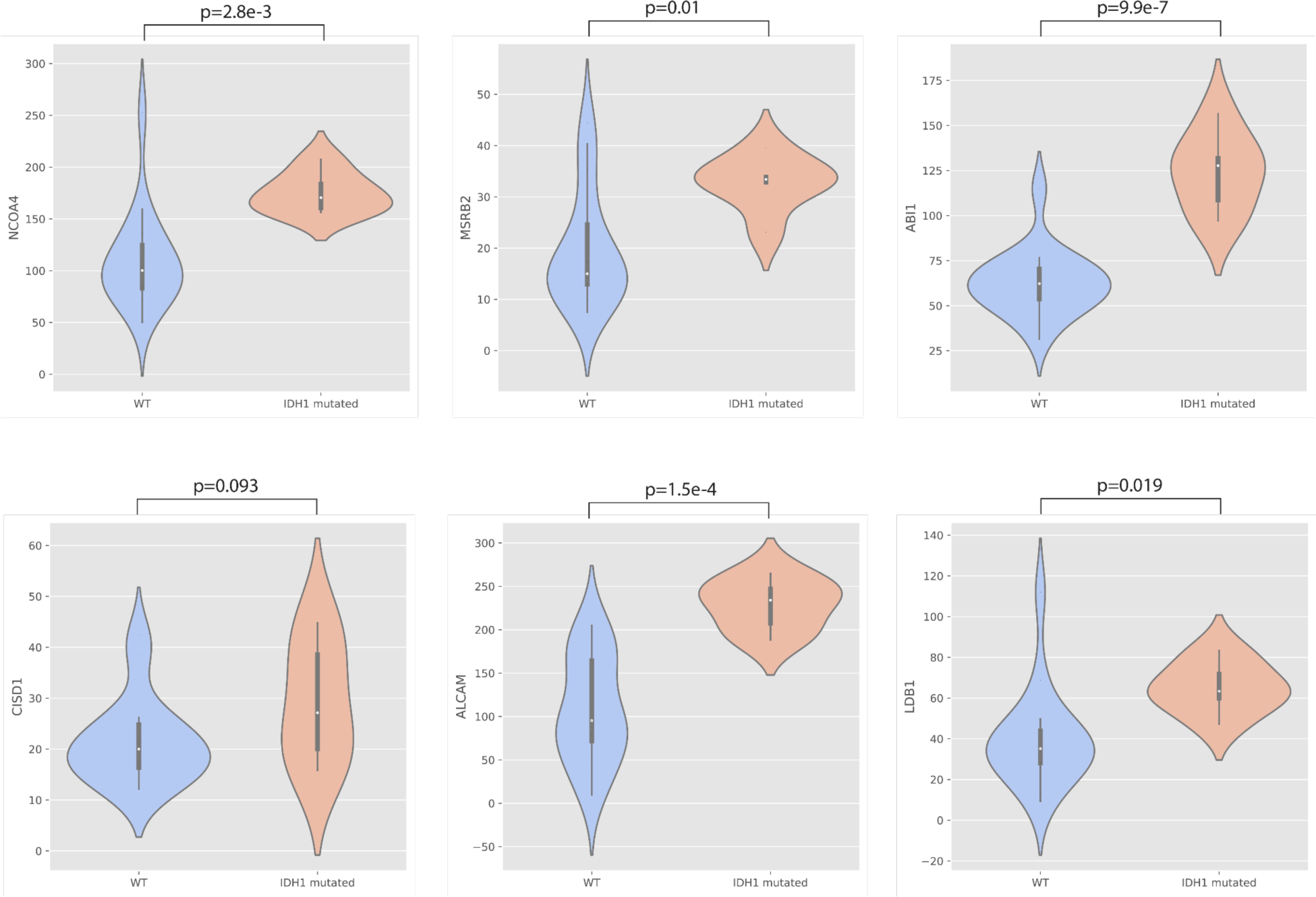
Data from Trautwein et al., supports that ETV7 regulon genes are upregulated in IDH1 mutants.

**Supplementary Figure 6:**
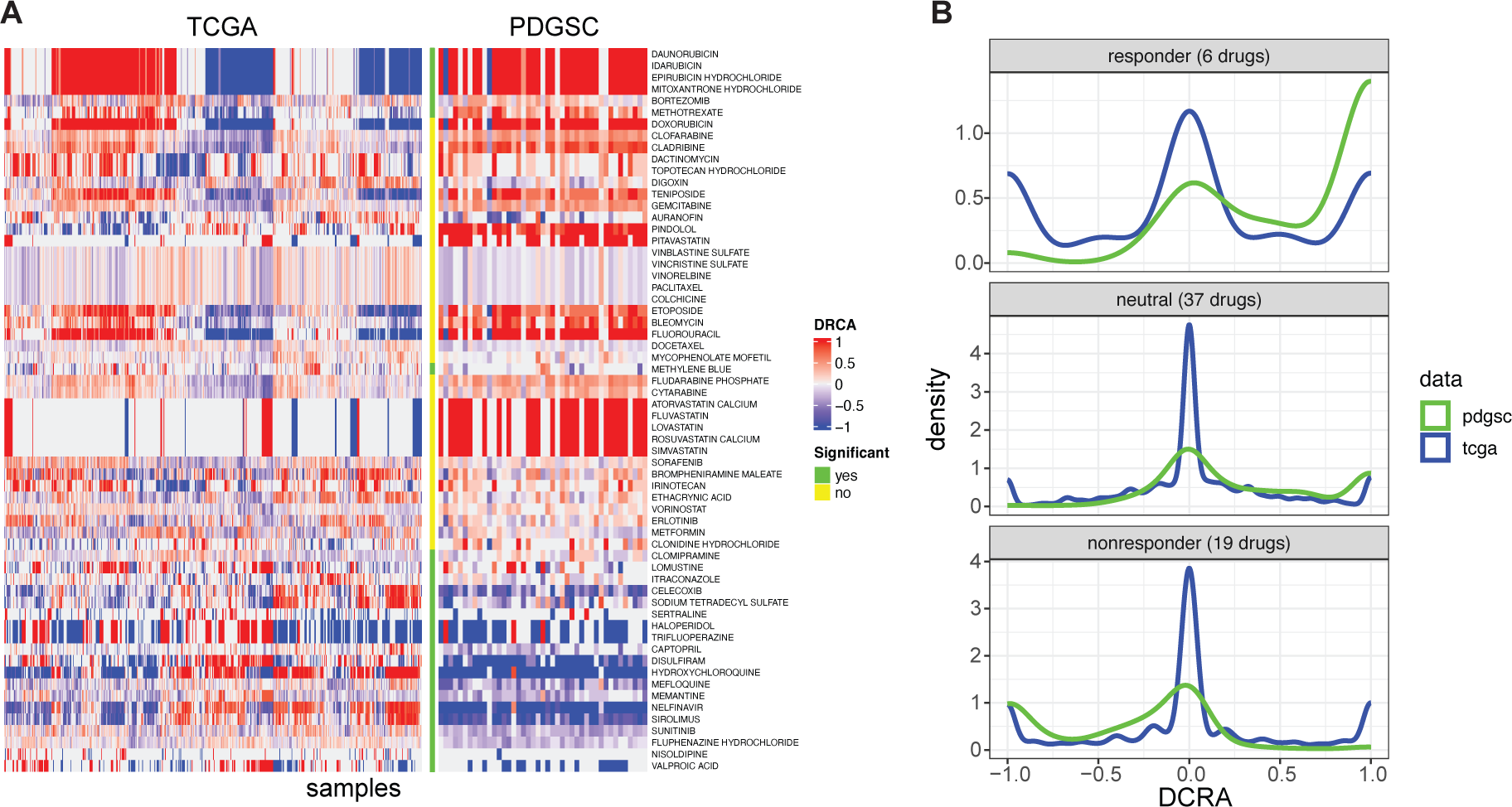
Comparison of PDGSC and TCGA training cohort drug constrained network activity (DC RA). **A)** Heatmaps of DCRA for both the TCGA and PDGSC samples for the drugs contained in the GBM drug screen. The drugs (rows) are organized for each drug based on the highest number of responder concurrencies from the top and number of highest nonresponder concurrencies from the bottom. The column between the two heatmaps identifies those drugs where gbmMINER was able to make statistically significant drug sensitivity predictions (>= 30 concurrencies based on an FDR of 0.05). The drugs were binned into 3 groups based on those drugs where gbmMINER correctly predicted at least either 30 responder classifications (top 6 rows), at least 30 nonresponder classifications (bottom 19 rows), and those in neither of the first two categories (middle 37 drugs). **B)** Density plots of the TCGA and PDGSC DCRA for the three binned groups of drugs (responders, nonresponders, and neutral). The PDGSC density for the responder bin is clearly skewed upwards in comparison to the TCGA density which explains why these PDGSCs were sensitive to these particular drugs. Similarly, the PDGSC density for the nonresponder samples was skewed lower than the TCGA density and thus shows why there was such poor sensitivity to those particular drugs. Alternatively, the neutral bin did not show much difference between the PDGSC and TCGA densities. Consequently, we did not observe a preponderance of concurrency skewed towards either responders or nonresponders.

**Supplementary Figure 7.**
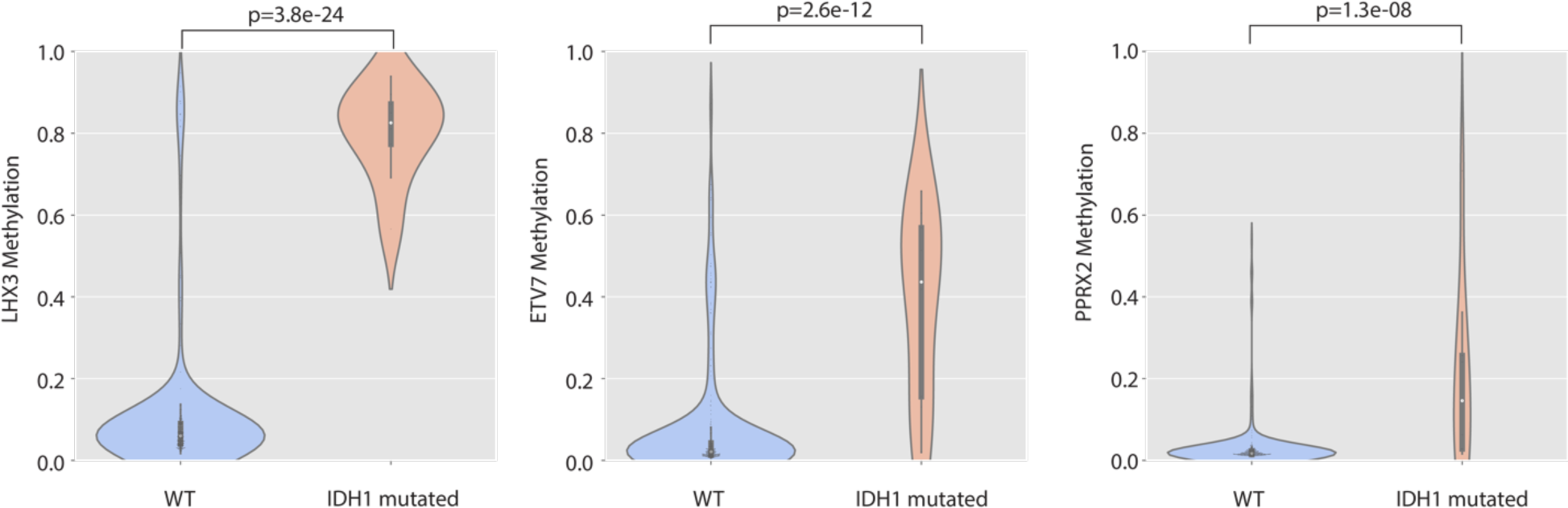
LHX3, ETV7 and PPRX2 have significantly higher methylation at their promoters in IDH1 mutants.

**Supplementary Figure 8.**
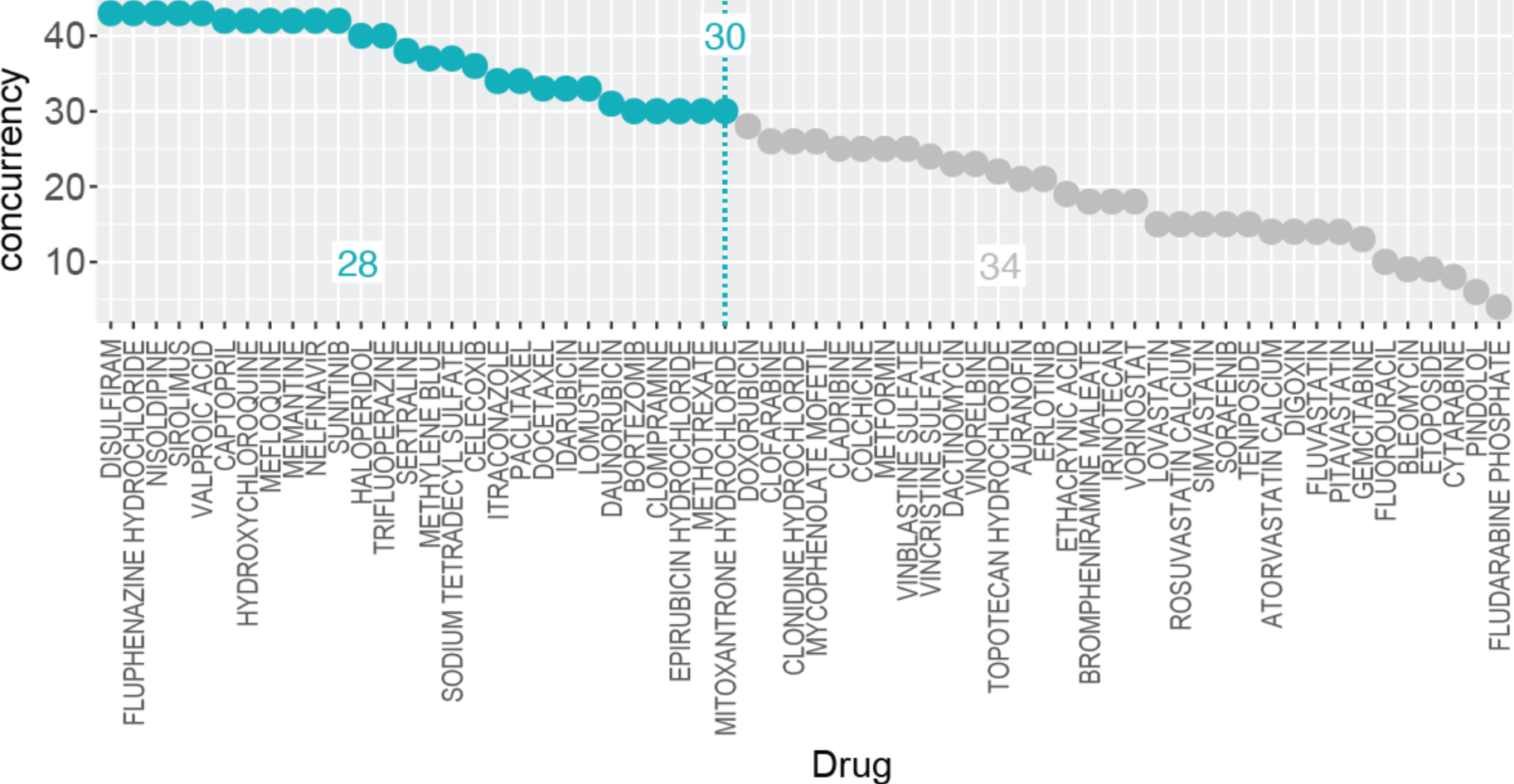
Distribution of the concurrencies for all the drugs tested. Green line marks number of concurrencies for the FDR < 0.05 and green dots indicate drugs with significant concurrencies.

**Supplementary Figure 9.**
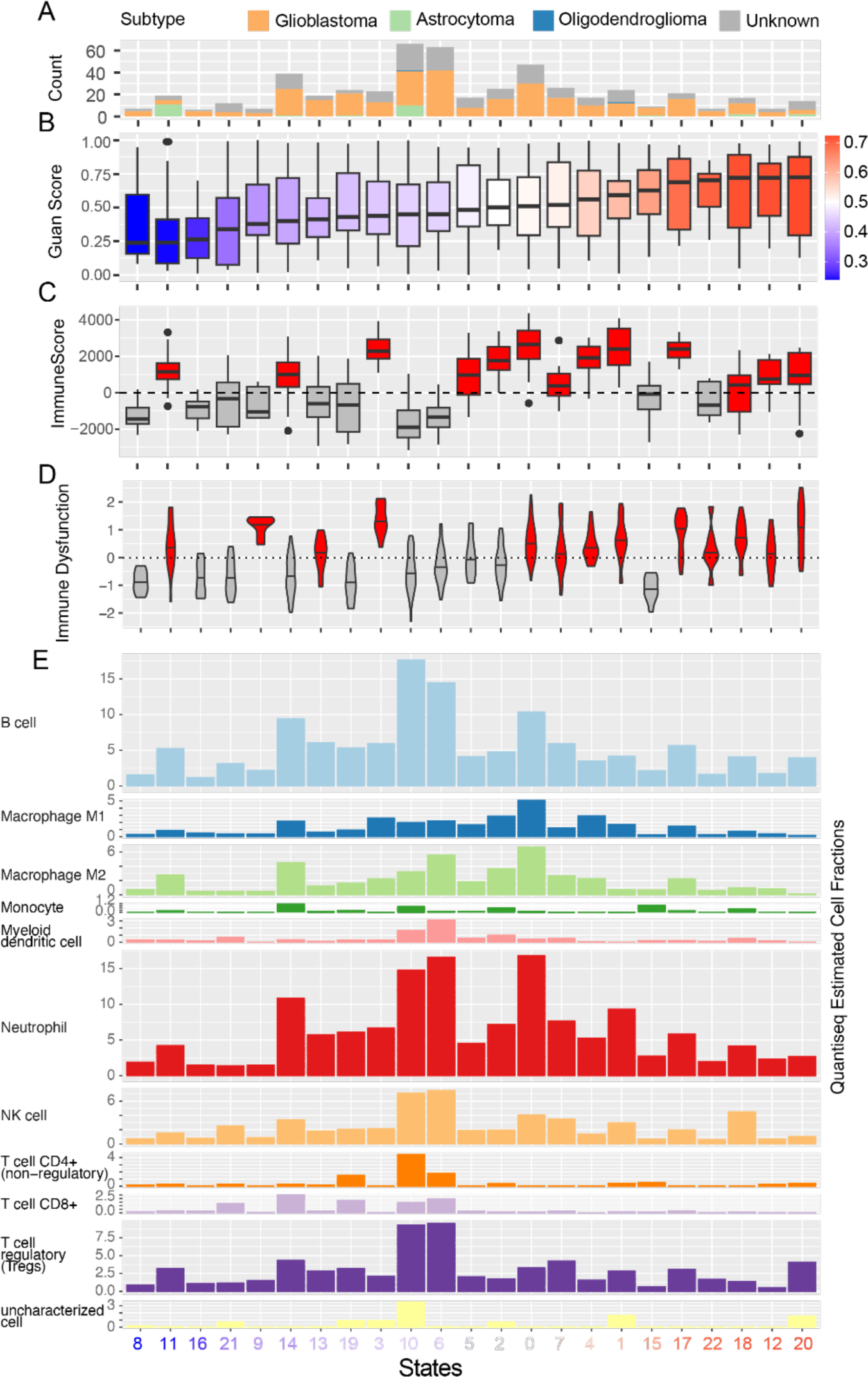
Distribution of Immune Dysfunction scores and immune cells across states. **A)** Distribution of the patient subtypes across each state identified by using 2021 WHO annotations provided by (Zakharova 2022). **B)** Boxplot of the transcriptional states rank ordered low to high risk from left to right, respectively) based on the distribution of the median Guan risk scores of patients included in the state. **C)** High risk states were associated with higher ImmuneScores indicating increased percentages of immune cell infiltration in the tumor. **D)** Higher risk states were with higher Immunescore are also associated with higher Immune Dysfunction score as determined by TIDE algorithm. E) Distribution of various immune cell types across states as determined by quantiSeq algorithm.

**Supplementary Figure 10.**
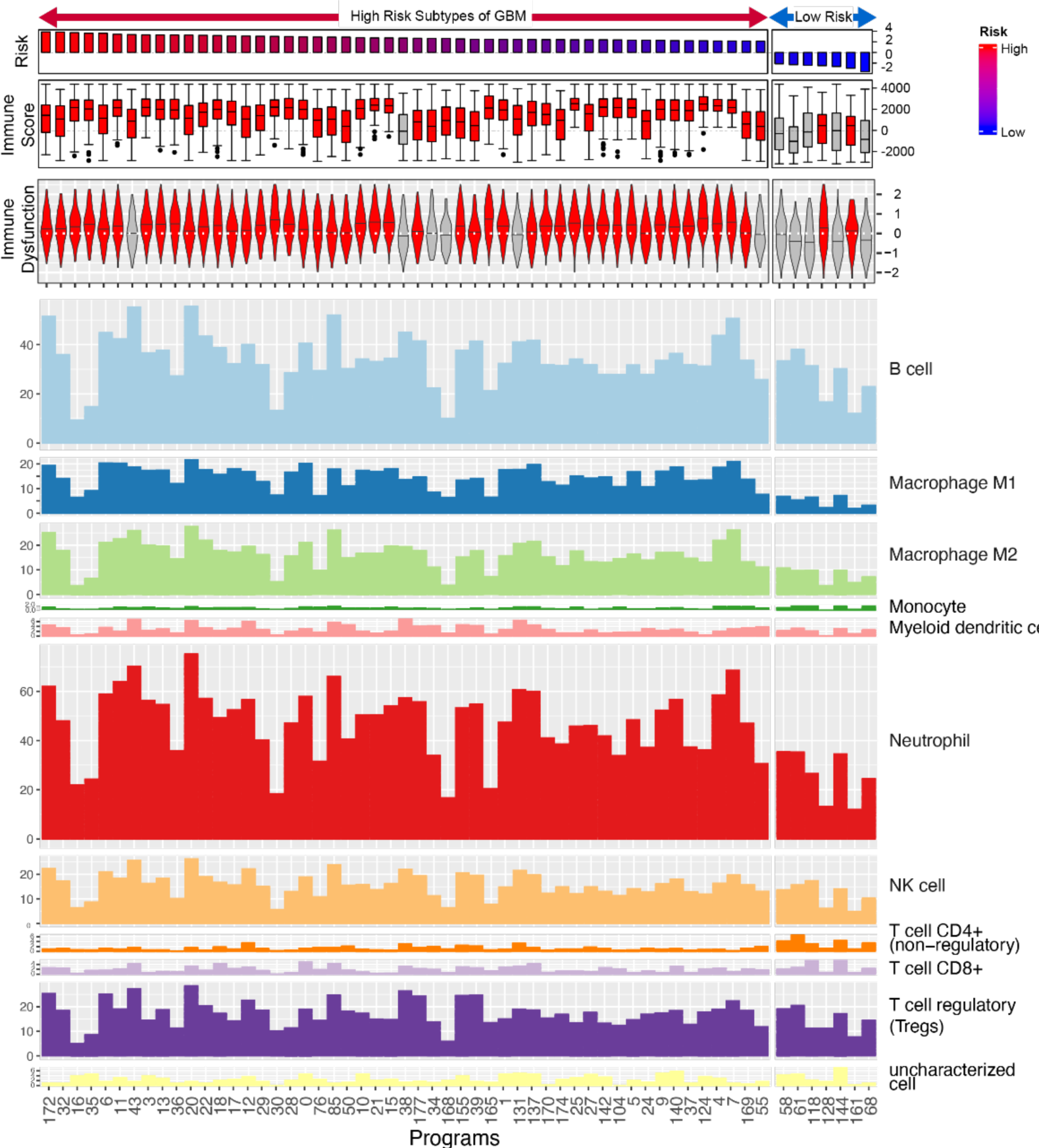
Distribution of Immune Dysfunction scores and immune cells across programs. **A)** Program risk scores, **B)** The level of immune cell infiltration for each program is indicated as a boxplot (red: median ImmuneScore > 0, gray: median ImmuneScore <= 0). **C)** The level of Immune Dysfunction as calculated by TIDE algorithm for each program is indicated as a violin plot (red: median Immune Dysfunction > 0, gray: median Immune Dysfunction <= 0). **D)** Distribution of main immune cell types across patients with overactive program activity.

**Supplementary Figure 11.**
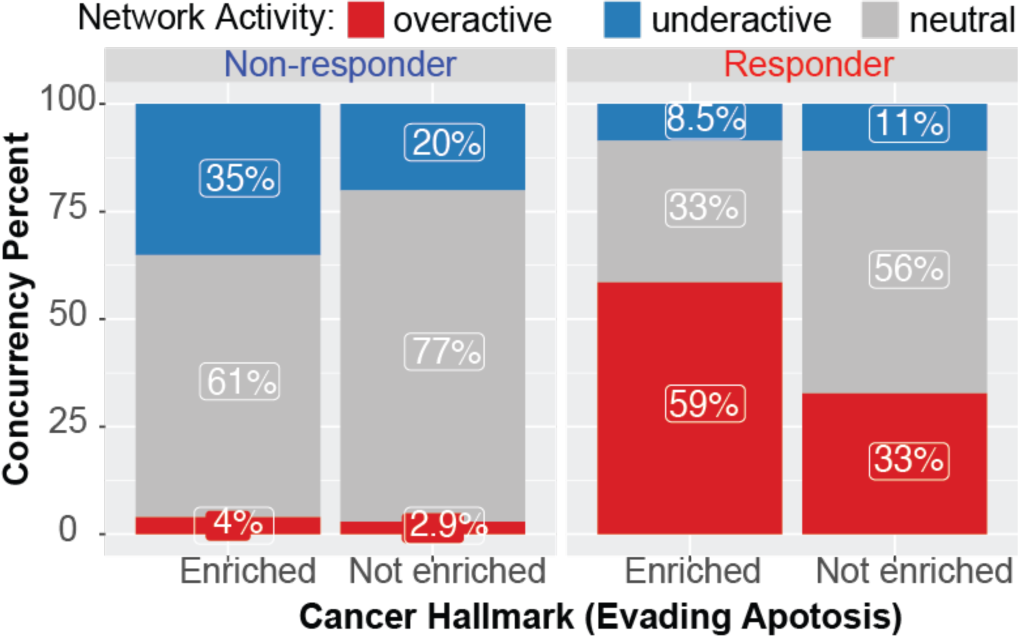
Activity status of “evading apoptosis”-enriched regulons containing targets of drugs that were concordant between DCRA-predicted and MTT-assay determined drug sensitivity across relevant responder and non-responder PD-GSCs. “Concurrency percent” refers to the proportion of concurrency events between predicted and observed drug sensitivity.

**Supplementary Figure 12.**
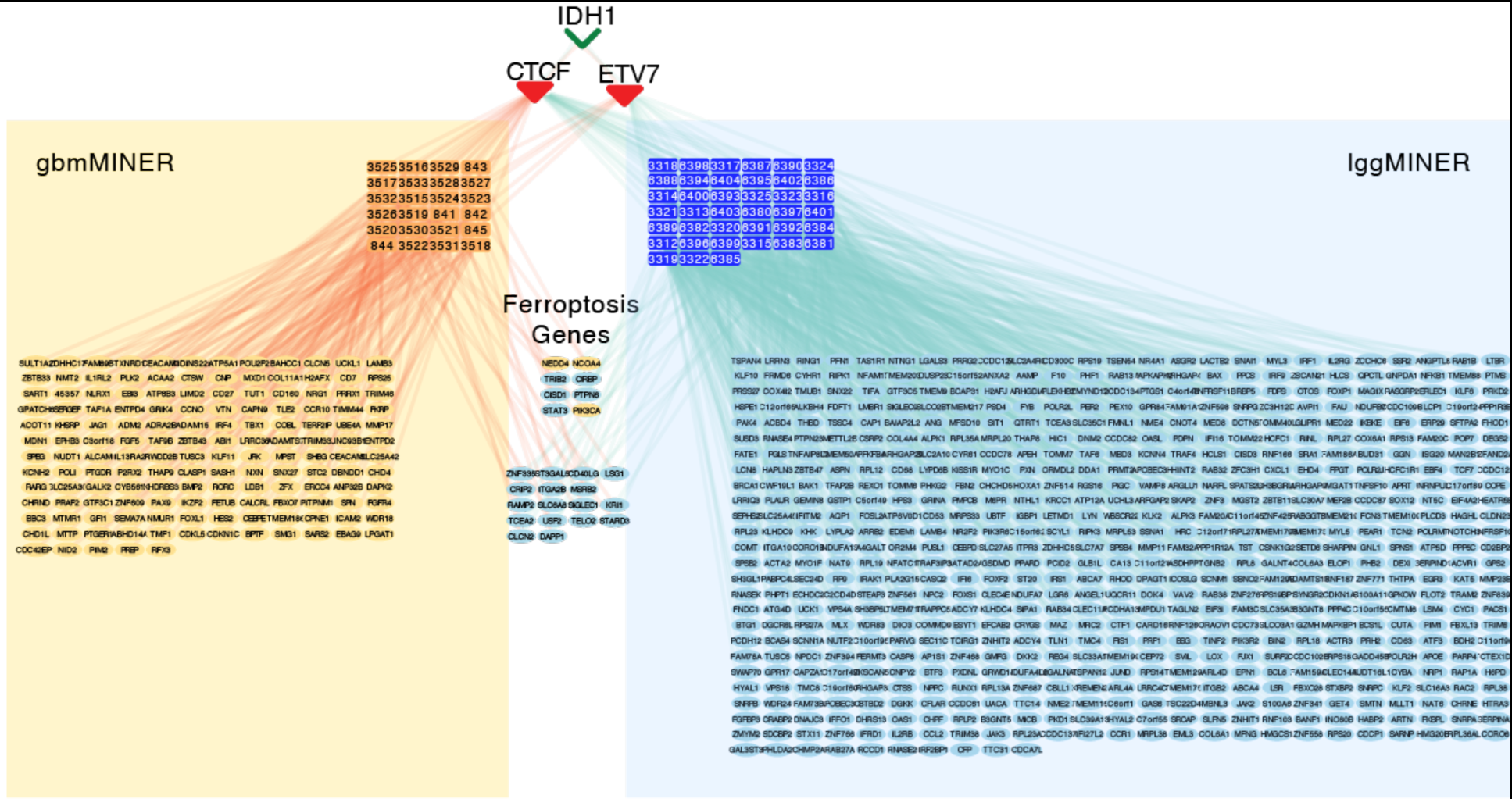
Overlap of gbmMINER and lggMINER network for ferroptosis related IDH mutation causal mechanistic flows.

