## Supplementary Table 5 for "An atlas of causal and mechanistic drivers of interpatient heterogeneity in glioma"

**Supplementary Table 5.** Known and new prognostic markers identified in gbmMINER

| **#** | **Mutated Gene** |  | **Regulators** | **gbmSYGNAL** | **gbmMINER** | **Regulons** | **Programs** | **Evidence** | **Reference** |
| --- | --- | --- | --- | --- | --- | --- | --- | --- | --- |
| 1 | MUC16 |  | HSF1, NFATC4, VSX1 | Yes | Yes | 3 | 3 | Mutated MUC16 is associated with better prognosis in Low Grade Gliomas but worst prognosis in GBM | (Ferrer 2023) |
| 2 | TTN |  | SOX10 | Yes | Yes | 1 | 2 | Hyper mutated in high Mutant‑allele tumor heterogeneity (MATH) group patients | (Wu et al. 2019) |
| 3 | OBSCN |  | PAX6, IRF7, IRF5, has-mir-338 | Yes | Yes | 4 | 4 | Hyper mutated in high Mutant‑allele tumor heterogeneity (MATH) group patients | (Wu et al. 2019) |
| 4 | PCLO |  | 12 | Yes | Yes | 17 | 11 | Patients with PCLO mutations showed poor prognosis | (Park et al. 2019) |
| 5 | AHNAK2 |  | 23 | Yes | Yes | 49 | 14 | All of the subclonal mutations of AHNAK2 were associated with 6poor prognosis in GBM. | (Bai et al. 2022) |
| 6 | SPTA1 |  | 11 | Yes | Yes | 16 | 11 | Mutated SPTA1 might be involved in GBM development | (Gao et al. 2019) |
| 7 | PKHD1 |  | ZNF652 | Yes | Yes | 1 | 1 | Mutations in PKHD1 are associated with poor prognosis in glioma patients | (Draaisma et al. 2015) |
| 8 | DNAH5 |  | 21 | Yes | Yes | 43 | 18 | All of the subclonal mutations of DNAH5 were associated with poor prognosis in GBM. | (Bai et al. 2022) |
| 9 | RYR2 |  | LHX1 | Yes | Yes | 1 | 1 | RYR2 was mutated in about 10% of the GBM patients | (Qin et al. 2020) |
| 10 | LRP2 |  | 36 | Yes | Yes | 57 | 22 | LRP2 mutations are associated with brain lower grade glioma | (Li et al. 2022) |
| 11 | MUC17 |  | 32 | Yes | Yes | 126 | 31 | Identified as potential biomarker for poor prognosis in adult low-grade glioma and glioblastoma patients | (Machado and Ferrer 2023) |
| 12 | CNTNAP2 |  | TFAP2C, SP1 | Yes | Yes | 2 | 2 | CNTNAP2 was one of the key genes with highest frequency of non-coding constraint mutations (NCCM) in GBM. | (Sakthikumar et al. 2020) |
| 13 | HMCN1 |  | 45 | Yes | Yes | 170 | 43 | Differential mutation analysis identified HMCN1 to be different between two risk groups | (Wang et al. 2022) |
| 14 | NF1 | Yes | 75 | Yes | Yes | 289 | 48 | NF1 is mutated in 53% of GBM mesenchymal subtypes | (D’Angelo et al. 2019) |
| 15 | RB1 | Yes | 10 | Yes | Yes | 11 | 6 | Mutations in RB1 was associated with proneural subtype | (Goldhoff et al. 2012) |
| 16 | PIK3CA | Yes | 14 | Yes | Yes | 15 | 12 | Missense mutations in PIK3CA promotes glioblastoma pathogenesis | (McNeill et al. 2018) |
| 17 | EGFR | Yes | 21 | Yes | Yes | 25 | 17 | Mutant EGFR enhances glioblastoma tumorigenicity by stimulating proliferation and inhibiting apoptosis. | (Nagane et al. 1996) |
| 18 | IDH1 | Yes | 133 | Yes | Yes | 442 | 77 | IDH1 mutation is sufficient to establish the glioma hypermethylator phenotype | (Lewandowska et al. 2014) |
| 19 | PIK3R1 | Yes | TCF3 | Yes | Yes | 1 | 1 | Somatic mutations in PIK3R1 act s oncogenic drivers | (Quayle et al. 2012) |
| 20 | PTEN | Yes | 14 | Yes | Yes | 19 | 8 | The most aggressive form of GBM has a very high frequency of mutation in the PTEN gene | (McLendon et al. 2008) |
| 21 | RELN |  | MEOX2, hsa-mir-129, hsa-mir-138 | No | Yes | 10 | 2 | RELN signaling was found to be associated with GBM pathology | (Schulze et al. 2018) |
| 22 | SYNE1 |  | ZNF410 | No | Yes | 1 | 1 | SYNE1 mutation is significantly correlated with the overexpression of several known GBM known markers | (Masica and Karchin 2011) |
| 23 | TP53 | Yes | 102 | No | Yes | 350 | 61 | The mutational status of TP53 is associated with GBM progression | (Krex et al. 2003) |
| 24 | ATRX | Yes | 108 | No | Yes | 387 | 70 | ATRX mutations are associated with IDH1 mutations | (Jiao et al. 2012) |
| 25 | KEL |  | POU3F3, GATA6, PARP1, NKX6-1, ETV7, TBX1, hsa-mir-222 | Yes | Yes | 9 | 6 | No known association to GBM |  |
| 26 | DNAH2 |  | REL, hsa-mir-9 | No | Yes | 7 | 2 | No known association to GBM |  |
| 27 | TCHH |  | PITX1, BRF1, CEBPZ, FOXD2, SPDEF, NFE2L2, SREBF2, hsa-mir-339, hsa-mir-500 | No | Yes | 8 | 5 | No known association to GBM |  |
| 28 | HSD17B7P2 |  | TEF | No | Yes | 1 | 1 | No known association to GBM |  |
| 29 | KRTAP4-11 |  | NR4A2, PAX6, ZNF652, POU3F4, EMX1, GATA6 | No | Yes | 6 | 5 | No known association to GBM |  |
| 30 | APOB |  | TBR1, ATF4, FOXC1, VSX1, CEBPB, XBP1, has-mir-375 | No | Yes | 11 | 6 | No known association to GBM |  |

*Known and new prognostic markers identified only in gbmSYGNAL*

| CDC27 |  | Yes | No |
| --- | --- | --- | --- |
| MLL3 |  | Yes | No |
| COL6A3 |  | Yes | No |
| POTEB |  | Yes | No |
| FLG |  | Yes | No |
| SCN9A |  | Yes | No |
| HEATR7B2 |  | Yes | No |
| TPTE2 |  | Yes | No |
| MUC2 |  | Yes | No |
| ZNF99 |  | Yes | No |
| HYDIN |  | Yes | No |
| TBC1D29 |  | Yes | No |
